# Integration of transcriptomics and long-read genomics prioritizes structural variants in rare disease

**DOI:** 10.1101/2024.03.22.24304565

**Authors:** Tanner D. Jensen, Bohan Ni, Chloe M. Reuter, John E. Gorzynski, Sarah Fazal, Devon Bonner, Rachel A. Ungar, Pagé C. Goddard, Archana Raja, Euan A. Ashley, Jonathan A. Bernstein, Stephan Zuchner, Undiagnosed Diseases Network, Michael D. Greicius, Stephen B. Montgomery, Michael C. Schatz, Matthew T. Wheeler, Alexis Battle

## Abstract

Rare structural variants (SVs) — insertions, deletions, and complex rearrangements — can cause Mendelian disease, yet they remain difficult to accurately detect and interpret. We sequenced and analyzed Oxford Nanopore long-read genomes of 68 individuals from the Undiagnosed Disease Network (UDN) with no previously identified diagnostic mutations from short-read sequencing. Using our optimized SV detection pipelines and 571 control long-read genomes, we detected 716 long-read rare (MAF < 0.01) SV alleles per genome on average, achieving a 2.4x increase from short-reads. To characterize the functional effects of rare SVs, we assessed their relationship with gene expression from blood or fibroblasts from the same individuals, and found that rare SVs overlapping enhancers were enriched (LOR = 0.46) near expression outliers. We also evaluated tandem repeat expansions (TREs) and found 14 rare TREs per genome; notably these TREs were also enriched near overexpression outliers. To prioritize candidate functional SVs, we developed Watershed-SV, a probabilistic model that integrates expression data with SV-specific genomic annotations, which significantly outperforms baseline models that don’t incorporate expression data. Watershed-SV identified a median of eight high-confidence functional SVs per UDN genome. Notably, this included compound heterozygous deletions in *FAM177A1* shared by two siblings, which were likely causal for a rare neurodevelopmental disorder. Our observations demonstrate the promise of integrating long-read sequencing with gene expression towards improving the prioritization of functional SVs and TREs in rare disease patients.

## INTRODUCTION

Long-read sequencing technology has dramatically improved in recent years in terms of accuracy and throughput[1,2]. This has unlocked a large reservoir of previously inaccessible variation, especially structural variation (SV), repeat expansions, and other complex variants [3,4]. In the clinical setting, however, exome and whole genome short-read sequencing (SRS) remain the dominant approaches to facilitate diagnoses of rare disease. While SRS increases the diagnostic yield of various rare diseases by 5 to 20% over exome sequencing, the diagnostic rates for rare diseases remain below 50%. While some fraction of these undiagnosed cases are likely due to non-Mendelian and/or non-genetic causes, the relatively low diagnostic rate underscores the need to explore long-read genome sequencing (LRS) as a new tool to increase the detection of pathogenic variants [5,6]. The potential clinical utility of LRS is especially apparent for rare SVs, which are known to have substantial impact on genome function [7], yet are systematically missed by SRS [3,4]. Furthermore, among rare SVs discovered by SRS, many variants’ impacts on genes are hard to interpret without additional functional validations, making rare disease diagnostic efforts difficult [8].

Gene expression data has been shown to help prioritize rare SNVs contributing to rare genetic disease, particularly using approaches that identify individuals with extreme expression compared to the rest of the population (“expression outliers”) [9–12]. Previous studies have shown a stronger enrichment of expression outliers having nearby rare SVs, suggesting a possible opportunity to prioritize rare functional SVs systematically [13,14]. Although numerous cases of rare SVs disrupting gene expression have been identified independently in the context of rare disease diagnostics, there is currently no systematic approach to effectively prioritize candidate disease SVs by integrating RNA-Seq and rare SVs genomic annotations. Notably, while STRVCTVRE [15], CADD-SV [16], and PhenoSV[17] can prioritize putative pathogenic SVs, STRVCTRVE only scores coding SVs, CADD-SV do not explicitly identify the affected gene and is trained to predict SVs under selection constraints but not regulatory SVs, and all tools have limited capability in finding functional SVs that modulate gene expression. In addition to SVs, variation at tandem repeat loci contributes to gene expression differences [18,19]. Repeat expansions at these loci have been implicated in a variety of rare neurological and neuromuscular diseases [20]. Because these expansions can potentially span thousands of base pairs, they are difficult to detect with SRS, but LRS methods have shown promise in detecting them [21].

In light of the current limitations regarding SV detection and assessment of functional impact, we propose two modalities for improving SV prioritization in rare disease diagnostics. First, we hypothesize that augmenting patient datasets with long-read sequencing will improve the detection of clinically informative rare and private SVs, large indels (30bp to 50bp) and complex variants where detection from SRS is more limited. Second, we recognize a major need to develop new methods that can prioritize rare SVs based on both genomic and functional -omics evidence, thereby providing better clues for clinicians to evaluate variants’ pathogenicity to rare diseases.

Addressing these needs, we performed long-read sequencing with Oxford Nanopore Technology (ONT) on 68 individuals from the Undiagnosed Disease Network (UDN) with pre-existing short-read genome sequence data to study the improvement in variant detection recall from SRS. Then, using blood or fibroblast RNA-Seq data from the same patients, we characterize the impact of LRS SVs on gene expression and investigate the utility of integrating these data to predict variant function. To address this challenge, we developed the Watershed-SV model and pipeline. Watershed-SV extends Watershed[9], a probabilistic variant prioritization model integrating multiple -omic signals and SNV genomic annotations, with several new SV-related annotation features to prioritize functional rare SVs. We trained and evaluated Watershed-SV models on GTEx expression outliers and SRS SV callset and found Watershed-SV outperforms the WGS-only baseline models in prioritizing coding and noncoding variants. When applied to the UDN patient dataset, Watershed-SV further provided additional disease-relevant SV candidates compared to CADD-SV, and provided direct insights into the gene disruption mechanisms.

## RESULTS

### Nanopore long-read sequencing of an exome-negative rare disease cohort

In order to assess the utility of long-read sequencing (LRS) and structural variant (SV) prioritization for rare disease diagnosis, we performed Oxford Nanopore (R9.4.1) LRS on 68 individuals from the Undiagnosed Disease Network (UDN) with a spectrum of clinical features who had inconclusive genetic testing using short-read sequencing (SRS) exome or genome (**Figure 1**). Across the cohort, we achieved a median sequencing throughput of 61Gb (range: 25Gb-133Gb), corresponding to an estimated aligned coverage of 18x (range: 7.1-33x) , and a median read length N50 of 19.2Kb (range 5kb-35kb) (**Figure S1a**). After read quality control, most samples had a median read quality score above 12.5 (range 9.9-15.1), corresponding to a median 93.6% read identity aligning to GRCh38 (**Figure S1b**).

**Figure 1.**
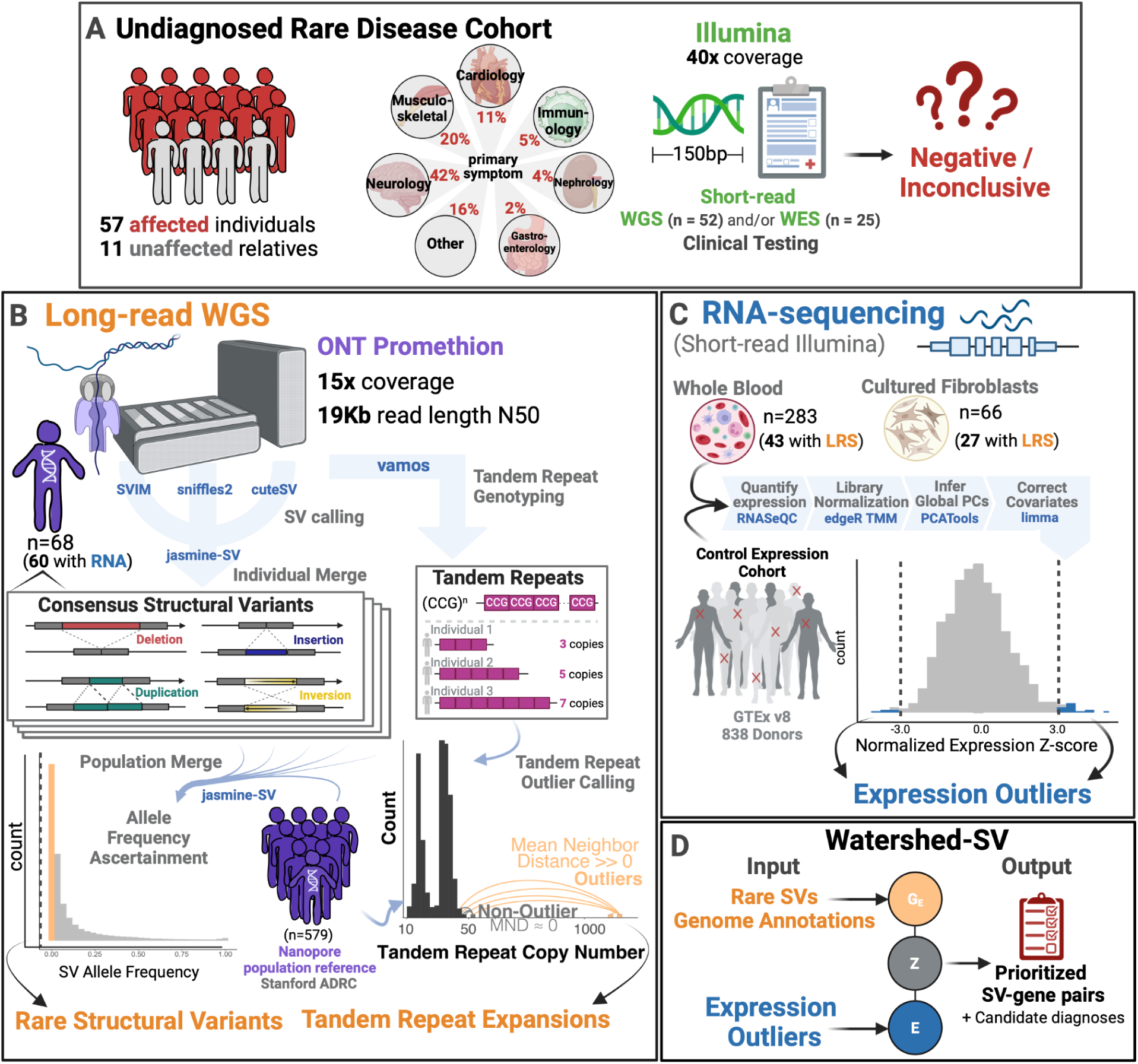
Undiagnosed patient cohort description and pipeline overview. **Cohort Description: A** Patients were recruited from the Undiagnosed Disease Network for a long read sequencing (LRS) study. These included 57 affected individuals and 11 unaffected family members from a wide range of primary symptom categories, including Neurology, musculoskeletal, and cardiology. Patients had previous short-read genetic testing with Illumina that was negative or inconclusive. **B Long-read Pipeline Overview**: individuals were sequenced on R9.4 flowcells on the ONT PromethION. Consensus structural variants were called by merging SVs across individual callers and keeping those that showed multi-algorithm support. A population merge of the UDN genomes together with Stanford ADRC population reference of 579 nanopore genomes, allowed ascertainment of robust allele frequencies for structural variants. Rare structural variants were filtered and intersected with overlapping genome annotations to input into Watershed. Vamos was used on a catalog of polymorphic tandem repeats to genotype tandem repeat copy numbers. A mean neighbor distance based outlier calling method was used to define extreme repeat expansions. **C** RNA-sequencing expression outlier pipeline: transcriptome data from the UDN was processed by quantifying expression, combining with tissue-matched controls from GTEx, normalizing for library size and composition bias, and correcting for batch effects and hidden factors. Expression outliers of the normalized data were input into Watershed. **D** Watershed-SV integrates signals from rare SVs and overlapping genome annotations to predict variants with large functional effects. High scoring watershed variants are prioritized and curated per patient for disease relevance.

### Long-reads detect more insertions and deletions than called from short reads

To compare the LRS genomes to the previously generated SRS genome data, we called structural variants (SVs) and large indels from both. Using a multi-algorithm consensus among sniffles2 [22], cuteSV [23], SVIM [24], facilitated by variant merging with Jasmine-SV [25], we detected 77,596 large indels (30bp to 50bp) and 120,950 SVs (>50bp) from LRS, 10-times and 2.7-times more, respectively, than were detected from standard clinical SRS SV calling with Illumina’s DRAGEN pipeline (**Figure 1b, 2a**). From LRS, we observed a balanced median number of 18,375 deletions (9,807 30-50bp, 8,788 50bp+) and 18,906 insertions (6,064 30-50bp, 12,815 50bp+) per individual, which is 3.9-times and 6.9-times respectively the median number identified by SRS (**Figure S1c**). We further detected a smaller median number of 436 duplications and 34 inversions per individual from LRS, while SRS detected more at a median number of 468 and 246 respectively, likely due to false positive calls from Manta-only SRS compared to confident consensus from LRS.

**Figure 2.**
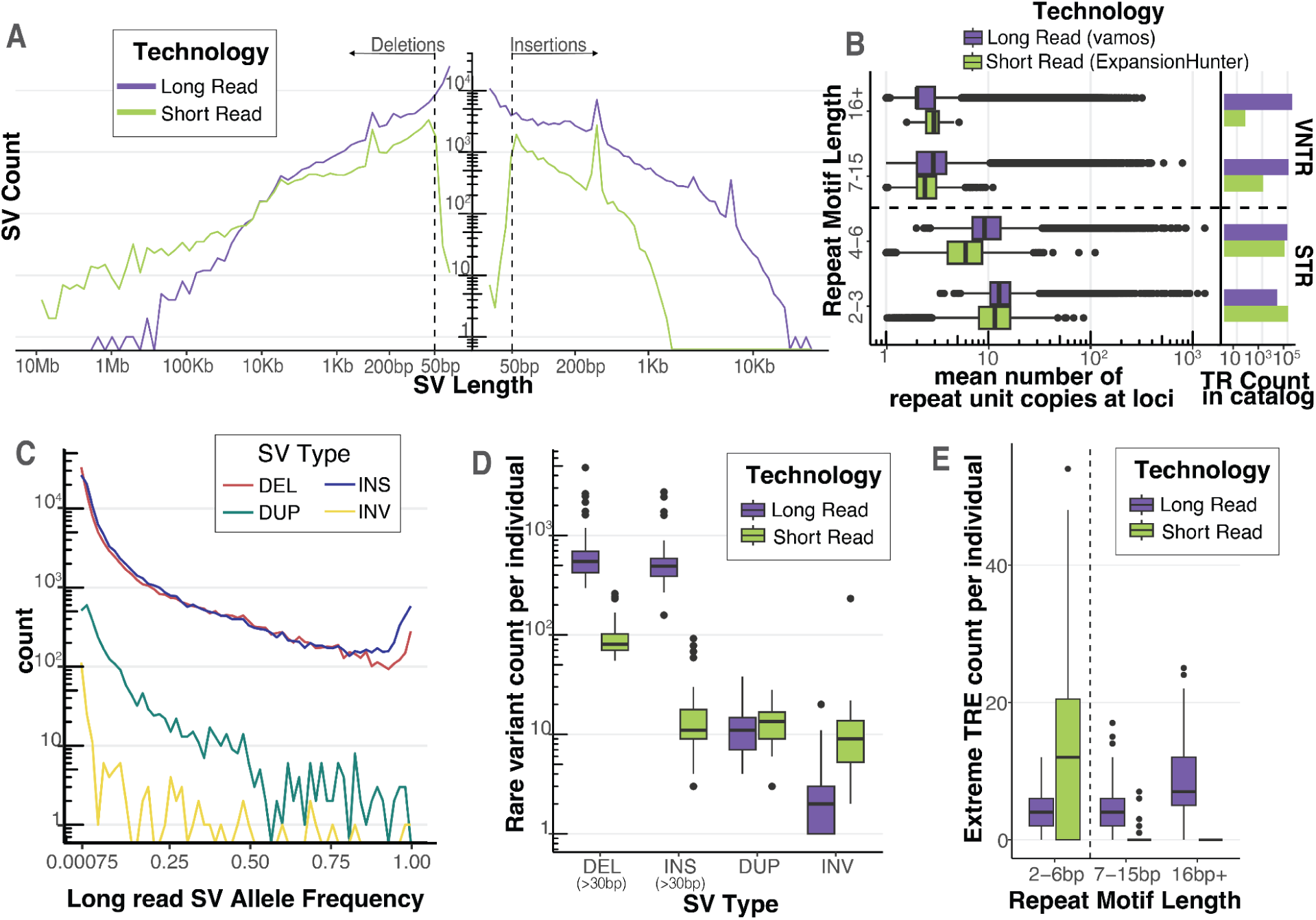
Long-read sequencing detects rare structural variants and extreme tandem repeat expansions. **A** Length distribution of deletions and insertions detected by each technology on a log-log axis. SVs were called with a consensus SV calling pipeline including SVIM, cuteSV, and sniffles2 for long reads and MantaSV calls were genotyped with paragraph for short reads. Dashed line represents 50bp, the threshold for calling an indel an SV. **B** Mean tandem repeat copy numbers estimated from the UDN genomes stratified by repeat motif length. Short tandem Repeats (STR) have repeat motifs between 2-6bp. Variable number tandem repeats (VNTR) have repeat motifs greater or equal to 7 bp. Vamos was used to genotype tandem repeat copy number in long reads and ExpansionHunter was used in short reads. Each tool used a different tandem repeat loci catalog to define TRs. Counts of TRs by repeat motif length bins present in the tools respective catalog is also plotted. **C** Allele frequency distribution of long-read discovered SVs from jasmine-SV merge with ADRC genomes. ADRC provided a reference sample of 600 nanopore genomes to allow robust estimation of minor allele frequencies. **D** Count of rare SVs (MAF < 0.01), detected per individual stratified by SV Type and Technology. Short read SVs were annotated with allele frequencies using SVAFotate and lookup in gnomAD, CCDG, and 1000G. **E** Count of extreme tandem repeat expansion (TRE) detected per individual. Extreme tandem repeat expansion outliers in each technology were called by jointly estimating repeat copy number distribution of long read vamos calls with the ADRC and of short-read ExpansionHunter calls with 1000G, and then calculating for each allele its average distance from its k nearest neighbors. Extreme TREs were defined as alleles with a standardized mean neighbor distance greater than 2, with k = 5 for long read and k = 25 for short read.

Stratifying by length, we see LRS identifies more variants in the 30bp to 50bp window for both insertions and deletions. LRS also identifies many more insertions across the entire length spectrum, particularly above 1kb, where SRS reads are too short to resolve any insertion sequence. For deletions, we also observe higher counts for LRS up to 10kb, after which we start to observe more variants from short reads (**Figure 2a**). This could be due in part to false positives in SRS callset or to alignment-based LRS methods not being calibrated to call deletions greater than 50kb. We find the length distribution of inversions and duplications to be roughly similar between LRS and SRS, though we do detect slightly more from short reads (**Figure S1d**).

### Long reads allow identification of longer, more complex variation at tandem repeat loci

Expansions of tandem repeats are known to cause multiple human diseases including myotonic dystrophy[26]. We tested the ability of long-read sequencing to resolve tandem repeat (TR) variation in the UDN cohort. To this end, we genotyped TR copy numbers with vamos [27] across 466,220 loci using LRS and ExpansionHunter across 174,260 loci using SRS [28] (**Figure 2b**). We used the recommended TR catalogs–a reference of where TRs exist in the genome and what the repeating motifs are–to run each tool and found they were widely characterizing different TRs; only 18% of loci were shared and, of those, 48% were annotated with different motifs (**Figure S2a**). Vamos defined their TR catalog from loci that varied in 32 haplotype-resolved PacBio HiFi genomes from the Human Genome Structural Variation Consortium and was designed to capture mostly complex variable number tandem repeats (VNTR) with motif lengths 7bp+, while the ExpansionHunter default catalog mainly captures short tandem repeats (STR) (2-6bp) that are small enough to be resolved with short reads. For the UDN cohort, we estimated the mean and variance of these TR copy numbers at each locus from both vamos (LRS) and ExpansionHunter (SRS) (**Table S2**). We discovered that vamos characterized loci that have higher mean TR copy numbers than ExpansionHunter, meaning LRS could detect longer repeats (**Figure 2b**). Further, the distribution of vamos TRs had a much longer tail for repeat motif length, mean repeat copy number, and repeat copy number variance than ExpansionHunter (**Figure S2c, S2d, S2e**). While almost all ExpansionHunter TRs had a mean repeat length of <100bp, vamos resolved TRs that had mean lengths well above 1kb (**Figure S2e**).

Direct comparison of LRS and SRS TRs can be impacted by their disparate TR catalog loci. To address this, we ran both tools using an identical TR catalog from STRchive (<u>harrietdashnow.com/STRchive</u>), a highly-curated set of 68 well-characterized and pathogenic TR loci. This allowed us to directly compare the LRS vamos and SRS ExpansionHunter on repeats with known disease-relevance. We calculated mean TR copy numbers across these 68 loci with both vamos and ExpansionHunter (**Table S3**). Vamos characterized all of the loci whereas ExpansionHunter only called 50 (73.5%) since some of these loci are longer than SRS fragment sizes. Of those called by short reads, we find a strong correlation across tools for the estimated mean TR copy number (Spearman correlation 0.833) (**Figure 2f**); however, there were a few repeats with larger discrepancies, most notably the CGC repeat in the gene XYLT1 that leads to the rare disease Desbuquois dysplasia-2 (ExpansionHunter mean=22, vamos mean=58), the AAGGG repeat in the RFC1 gene that leads to Cerebellar ataxia with neuropathy and vestibular areflexia syndrome (ExpansionHunter mean=16, vamos mean=37), and the CCTG repeat in *CNBP* that leads to Myotonic dystrophy type 2 (ExpansionHunter mean=17, vamos mean=38) (**Table S3**). As vamos was developed for noisier long-read data, it is more permissive of assigning non-perfectly matching repeats to the input motif, particularly in nested repeats where the pathogenic motif is flanked by others at the same locus, which may explain why vamos calls higher repeat copy number means compared to ExpansionHunter. For each repeat locus, the correlation across individual haplotypes was more variable (median Spearman’s correlation = 0.635, range= 0.163 – 0.850) (**Figure S2g**).

### Long-read population references enable frequency estimates for rare SVs and TREs

For rare disease diagnosis, it is critical to determine which variants are rare in the population. For LRS SV this is a major challenge as existing short-read references such as gnomAD, CCDG, and 1000G, show low ascertainment of SVs discovered from LRS data (**Figure S1f**) [29–31]. To address this issue, we used a technology-matched population reference of nanopore genomes generated from Stanford’s Alzheimer’s Disease Research Center (ADRC) and the Stanford Aging and Memory Study (SAMS) (n=571) [32]. ADRC + SAMS nanopore genomes were processed with the same SV calling workflow and were merged using Jasmine-SV with the UDN callset; allele frequencies were estimated based on merged allele counts in the combined set. We observe the expected allele frequency distribution for SVs, finding 32.6% of variants are rare (minor allele frequency < 0.01) (**Figure 2c**) (**Table S4**). For SRS SVs, we observed a high genotyping rate of our Manta callsets with Paragraph and used them to determine the allele frequencies for SRS SV calls (Supplemental Figure 1E). We filtered both LRS and SRS SVs down to rare variants with MAF < 0.01 and found each genome had a median burden of about 546 rare deletions and 490 rare insertions detected by LRS, whereas by SRS methods there was a median burden of 80.5 rare deletions and 11 rare insertions (**Figure 2d**). We further observed a smaller burden of rare duplications and inversions at a median number of 11 and 2 respectively from LRS, and 13.5 and 9 in SRS (**Figure 2d**).

Using the combined population of over 600 genomes of our UDN and ADRC + SAMS cohort [32], we dramatically improved our ability to filter LRS SVs for rare disease diagnosis. We detected a median of 716 rare SVs (>50bp) per genome, whereas using a short-read references (i.e. gnomAD) to ascertain rare variants, we would have detected over 17,000 per genome, making curation of these variants extremely difficult (**Figure 1g**). Genotyping LRS SVs in 1000G short-read genomes with PARAGRAPH to ascertain allele frequencies yielded a median of 3,198 SVs per individual, still 4.4 times more than we discover from a technology-matched reference (**Figure S1g**).

Similarly, we sought to assess TR distribution to identify rare tandem repeat expansions (TREs) in the UDN cohort, that is, extreme outliers of TR copy number. Using the ADRC reference for long-read sequencing (LRS), we developed an algorithm to call rare TREs by thresholding on a standardized mean neighbor distance (MND) (see **Methods)**. This method evaluates how much longer on average an allele is from its K-nearest neighbors in the repeat copy number distribution. This statistic is designed to distinguish clearly separated and expanded alleles from those that are just at the tail of the distribution, enabling us to identify samples with rare repeat expansions (see schematic in **Figure 1b**). We apply this method to vamos TR genotypes . Setting K = 0.5% of total allele number, we found a median of 14 TREs per individual, in comparison to median of 427 using Tukey’s outlier approach based on empirical TR copy number distribution, greatly reducing the false positive TRE calls.

To compare the characteristics of LRS and SRS TREs, we additionally assessed allele frequency and identified rare TREs from SRS with MND using genome-wide repeat catalog of 1000G as a reference. While ExpansionHunter detects more rare TREs from SRS that are STRs (2-5 bp) (median EH = 22, median vamos = 8), vamos detected more rare variable number tandem repeats (VNTR, 7+bp) TREs from LRS (median EH = 0, median vamos = 11). Further, vamos TREs were in loci with longer motif unit lengths (median = 22) compared to ExpansionHunter (median = 2) (**Figure S3c**). ExpansionHunter and vamos rare TREs had similar distributions of standardized MNDs, yet vamos was able to find outliers in repeats with longer mean lengths, and the expansion allele lengths were much larger in the LRS TREs compared to SRS data (**Figure S3d, S3e, S3f**).

### Transcriptome data identifies functional effects of rare SVs

To characterize the functional effects of rare structural variants (SV) and extreme tandem repeat expansions (TRE) detected by LRS, we used RNA-seq obtained from individuals in our UDN cohort. Most (n=60) of the UDN LRS subset also had short-read RNA-seq data generated as part of their UDN enrollment, including (n=42) samples from whole blood and (n=27) from fibroblasts. We hypothesized that impactful SVs would drive outlier expression of nearby genes. We called gene expression outliers from blood and fibroblast tissues by jointly normalizing the UDN cohort with matched-tissue RNA-seq samples from GTEx ,and adjusting for RIN, sex, batch, and global expression principal components as covariates, and observed a median of 139 outliers per individual in UDN blood samples (**Figure S4a)**. We found strong enrichment at a log odds-ratio (LOR) of 0.63 (adj. p-value = 4.8e-6) for rare SVs nearby (within 10kb) of expression outlier genes. We observed enrichment across all types of SVs, with stronger enrichments for more stringent z-score thresholds defining outliers (**Figure 3a**). These observations are concordant with previous analysis in GTEx, where CNVs were enriched for proximity to genes with outlying expression [9,13].

**Figure 3.**
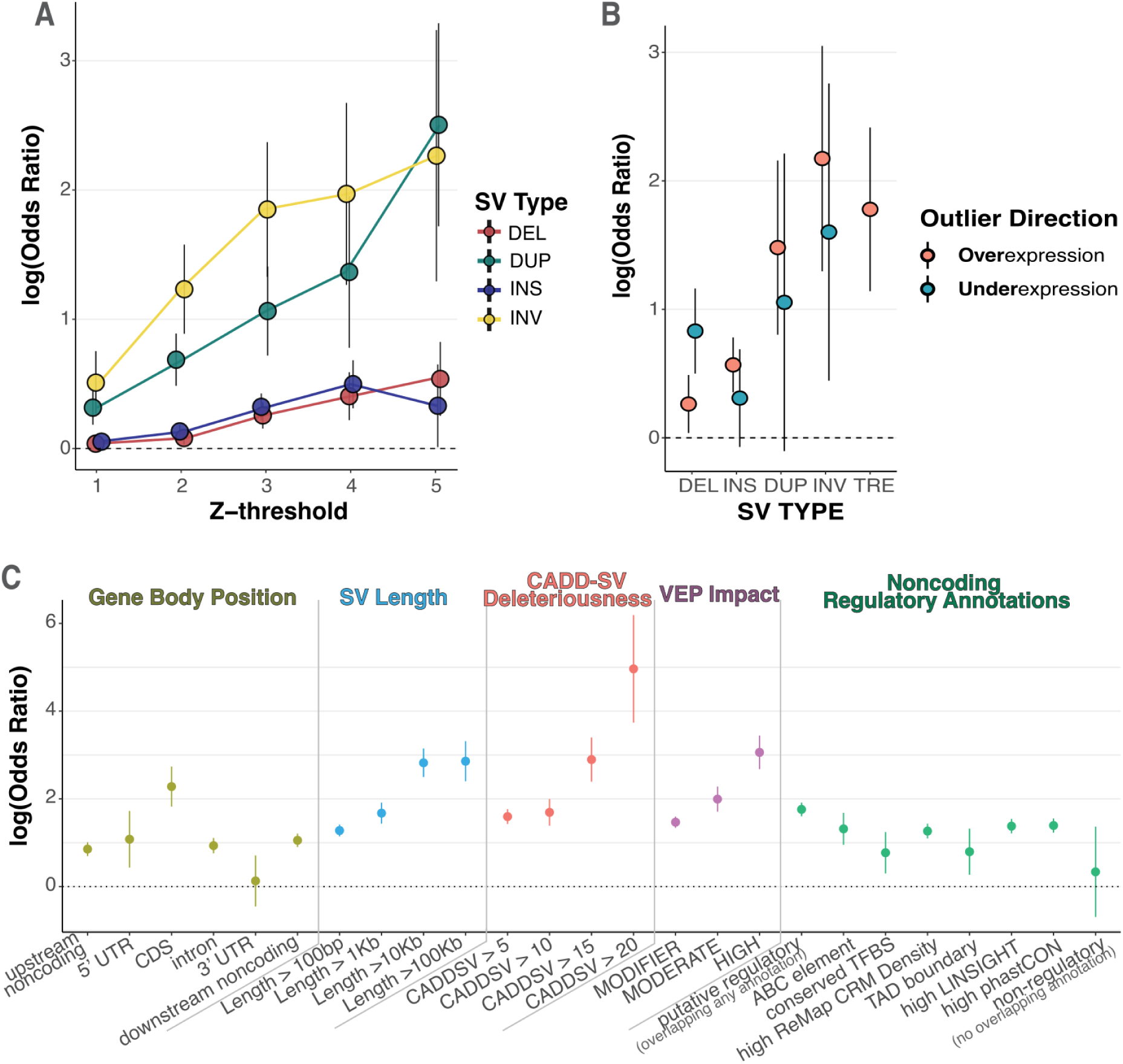
Rare long-read-discovered SVs are strongly enriched proximal to gene expression outliers. **A** enrichment of rare structural variants, stratified by type, within 10kb of an expression outlier gene given the specified absolute Z-score threshold. Estimate of log odds ratio plotted with error bars representing standard errors of the estimate. **B** Directional enrichment for rare SVs within 10kb of either over (Z > 4) or under (Z < -4) expression outliers. Model for tandem repeat expansion (TRE) nearby underexpression outliers did not converge due to the lack of examples of rare TREs near underexpression outliers, so was not be plotted. **C** Enrichment of rare SVs, across all SV types, within 100kb of expression outliers, stratified by genome and variant annotation categories. Gene body position displays enrichment of VEP annotated categories for SV location relative to the gene body of the expressed gene. If an SV overlaps multiple categories it is assigned to the one with highest priority given the following ordering: CDS, 5’UTR, 3’UTR, intron, upstream noncoding, downstream noncoding. SV length and CADDSV deleteriousness display enrichment of rare SVs with length and CADDSV score respectively above the specified threshold. VEP impact displays enrichment of rare SVs with the given VEP impact category, where HIGH represents predicted loss-of-function variants. Finally, we display enrichment of SVs that overlap with noncoding regulatory annotations, including if it overlaps an ABC regulatory element linked to the expressed gene, a conserved transcription factor binding site (TFBS), a high-density of ChIP-seq peaks defining conserved regulatory modules (CRM) from ReMap, a TAD boundary detected in multiple cell types, highly constrained LINSIGHT SNVs, or a highly conserved region by phastCON. We also display enrichments for SVs that overlapped any one of these annotations (putative regulatory SVs) and for SVs that do not overlap with any of these annotations (putative non-regulatory).

Using the improved variant discovery enabled by LRS, we expand upon previous results by analyzing insertions and TREs, which are difficult to call with SRS. We observed that genes from individuals with outlier expressions are significantly enriched for nearby rare insertions and TREs (z-score threshold = 4, **Insertion** LOR (95% CI) = 0.496 (0.13-0.86), **TRE** 1.72 (0.47-3.0))(**Figure S4b)**. We observed the strongest enrichment for duplications and inversions, which may be due to the fact that these variants are on average much longer (INV mean = 103kb, DUP mean = 18.4kb) than deletions and insertions (DEL mean = 450, INS mean = 275). Stratifying enrichments by outlier direction, we also observed that deletions were enriched for under-expression events (LOR (95% CI) = 0.83 (0.18-1.48)), and insertions, duplications, and TREs were enriched for overexpression events (**insertion** 0.568 (0.149-0.987), **duplications** 1.48 (0.153-2.81), **TRE** 1.78 (0.53-3.01)). Inversions were enriched for outliers in both directions, suggesting that large inversions may rearrange regulatory landscapes to increase or decrease expression of nearby genes (**Figure 3b**).

Previous work in GTEx based on SRS did not report significant expression outlier enrichments for insertion variants, which may be a reflection of the limited power of SRS to call insertions[9]. To characterize differences in enrichments based on sequencing technology, we compared LRS enrichment to those calculated based on the UDN SRS SVs. We observed the LRS callsets had larger enrichments than SRS callsets, particularly for insertions where SRS insertions were not enriched similar to what was previously observed in GTEx, underscoring the utility of LRS for discovery of novel functional insertions (**Figure S4c**). When considering other variant types, we observed that most rare SVs near outlier genes were detected uniquely by LRS (77.3% of deletions and 97.2% of insertions). Rare duplications found nearby outliers were largely detected by both technologies (57.1% shared). In contrast, rare inversions nearby outliers were frequently only detected by short-reads (71.4%), but these variants had only modest enrichments, suggesting a higher false discovery rate of inversions from SRS (**Figure S4d**).

To characterize rare variant enrichments near expression outliers, we estimated SV enrichments stratified by several annotations, increasing our proximity-window size to 100kb to capture more non-coding variants. Among gene body regions, we observed the strongest enrichment for SVs overlapping protein-coding sequence (LOR 2.28, adj. p-value = 1.5e-05). We also observed enrichments for non-coding SVs, including intronic (LOR 0.93, adj. p-value = 2.6e-06), upstream noncoding (LOR 0.85, adj. p-value = 1.6e-06) and downstream noncoding regions (LOR 1.05, adj. p-value = 1.5e-10). As the length of the SV increased, we observed enrichments up to log odds ratios of 2.86 (adj. p-value = 1.2e-08) for variants longer than 100kb, indicating that variants that disrupt more sequence are more likely to drive outlier expression. Further, increasing thresholds for variant deleteriousness as predicted by CADD-SV [16] or VEP impact categories [33] was also associated with increased enrichment. Variants with high CADD-SV score (>20) were strikingly enriched at a log odds ratio of 4.96 (adj. p-value = 1.3e-03) (**Figure 3c**).

We then estimated enrichment for non-coding rare SVs using several regulatory annotations. We included the proximity to a coding SV as a covariate in all enrichment analyses to mitigate effects due to tagging of coding SVs. We observed that SVs that intersect an activity-by-contact (ABC) mapped regulatory element [34] were enriched for outlier expression of the ABC target gene (LOR 1.32, adj. p-value = 7.7e-03). Rare SVs were also enriched for outliers when overlapping a high density of ChIP-seq peaks from ReMap [35] (LOR 1.27, adj. p-value = 1.1e-12), noncoding regions with evidence of negative selection from LINSIGHT variants [36] (LOR 1.38, aj. p-value = 1.6e-15), or highly conserved sequences by phastCons [37] (LOR 1.39, adj. p-value = 2.2e-16). As a negative control, we curated a set of putatively non-regulatory SVs, non-coding SVs that did not overlap with any of the previous annotations or any conserved transcription factor binding motifs, TAD boundaries, ENCODE conserved cis-regulatory elements, or high scoring CADD variants. These 519 non-regulatory rare SVs were not enriched proximal to expression outliers (LOR 0.33, adj. p-value = 1.00) (**Figure 3c**). We then sought to validate these ABC-element enrichments in a larger sample size from GTEx. Notably, enhancer enrichments were observed for single-tissue outliers but not multi-tissue outliers (**Figure S4e**).

### Watershed-SV integrates genomic annotations and transcriptomic signals to prioritize rare functional SVs

The co-occurence of expression outliers with rare SVs provides the opportunity to jointly use genomic annotations and transcriptomic signals to prioritize rare functional SVs. To do so, we extended our method Watershed [9], originally created for SNVs, to develop Watershed-SV to assess structural variants. Watershed-SV models individual gene expression jointly with genomic annotations of nearby SVs to infer the probability the individual harbors a high-impact regulatory SV (“Watershed posterior probability”). We included annotations characterizing the gene-body and separate annotations for flanking sequence (up to either 10kb or 100kb from the gene body). We then trained the Watershed-SV models using individual-gene pairs from the GTEx v8 SRS SV callset. We first developed three Watershed-SV models for gene expression in skeletal muscle, whole blood, and cell cultured fibroblasts tissues, which we refer to as tissue-specific Watershed-SV models. We then developed a multi-tissue Watershed-SV model using expression outliers defined from the median expression z-score across 48 GTEx tissues. To assess the performance of the Watershed-SV models, we evaluated expression outlier predictions of the model using held-out pairs of individuals who share the same rare SV genotype near a particular gene (“N2-pairs”). We used the Watershed-SV posterior probability from one sample in the N2-pair as a prediction and the expression outlier status of the genotype-matched, held-out sample as the label. We compared the Watershed-SV models to a model that included genomic annotations but no expression information (“WGS-only baseline”). The 10kb multi-tissue Watershed-SV model outperformed the WGS-only baseline (ΔAUPRC bound = (0.13, 0.27)) (**Figure 4a**), demonstrating the utility of incorporating gene expression information in SV prioritization. Watershed-SV learned large feature importance scores for SV length, overlapping with exons, location in 5’ UTRs, CADD scores, and for disruption of cis-regulatory elements. (**Figure 4b, Figure S6**).

**Figure 4.**
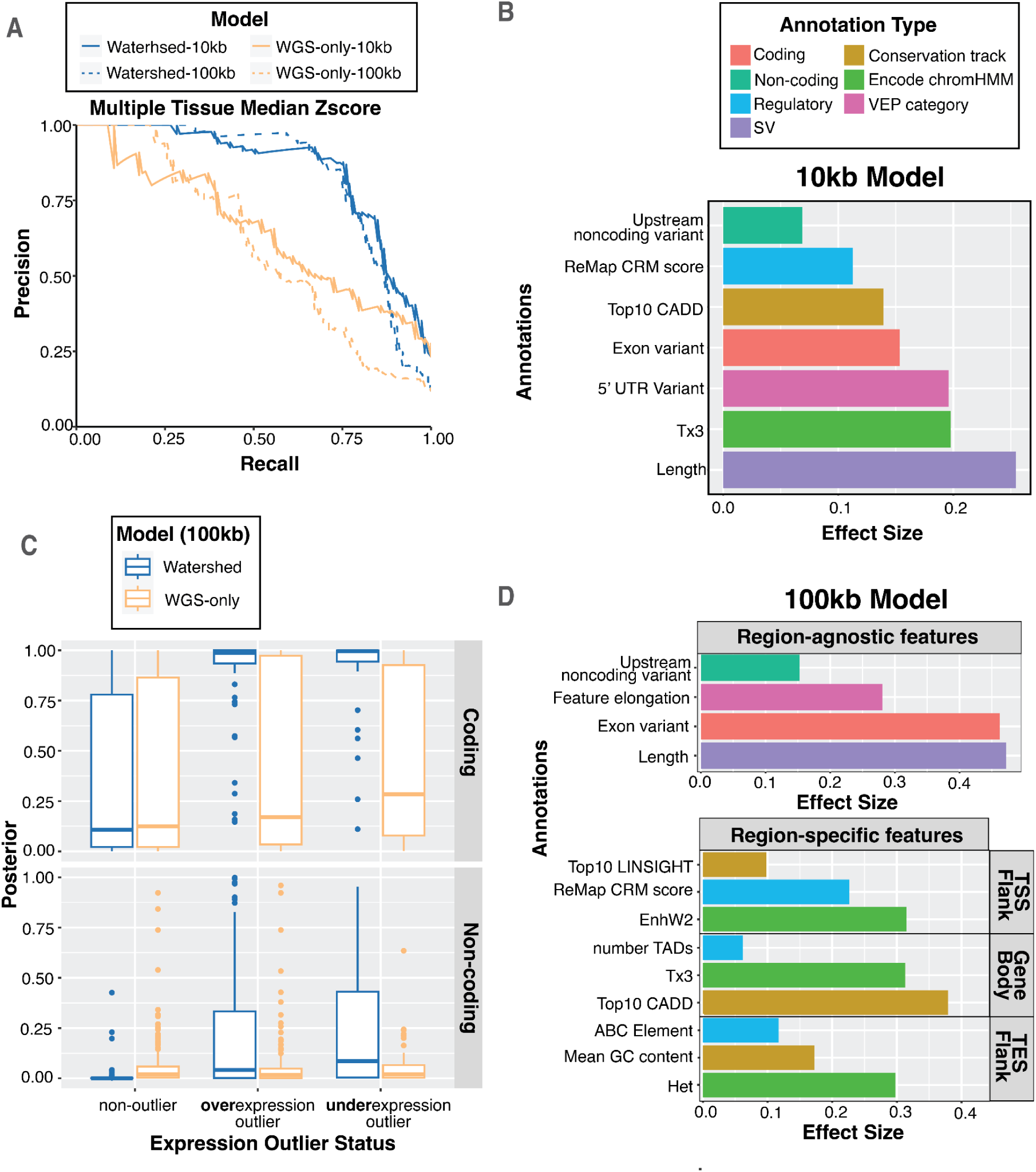
Watershed-SV improves prioritization of rare SVs in healthy and muscular dystrophy cohort. **A** Precision-Recall Curves (PRC) of benchmark using held-out N2 pairs; We ran multi-tissue Watershed-SV using both 10kb (solid) and 100kb (dashed) distance limit as well as WGS-only model with the same setup. **B** top positive genomic annotation effect sizes (β) for 7 major categories of the 10kb multi-tissue Watershed-SV model. **C** Using a z-score threshold of -3 and 3, we stratified 100kb multi-tissue Watershed-SV model prediction on CMG muscular disorder dataset posterior probabilities by under-, over-, and non-outliers (column), and then by coding vs noncoding variants (row); each dot represent an gene-SV pair. **D** top positive genomic annotation effect sizes for 100kb multi-tissue Watershed-SV model. 7 annotation categories are grouped into region-specific (TSS/upstream Flank, Gene Body, TES/downstream Flank) and region-agnostic features. Region specific features are separately aggregated for each SV, then collapsed to each gene by regions.

To capture longer range SV-gene interactions in the 100kb Watershed-SV model, we included ABC [34] enhancer-gene links among the annotations. We categorized annotations into three categories based on SV location: upstream flanking region (UFR) annotations, gene body annotations, and downstream flanking region (DFR) annotations. Watershed-SV learned especially large weights for UFR annotations, reflecting the known functional importance of regulatory elements near promoters. Among gene body annotations, we observed a high effect size CADD scores and active transcription annotations (**Figure 4d, Figure S7**). The 100kb multi-tissue Watershed model also outperformed the baseline WGS-only model (ΔAUPRC bound = (0.12, 0.26)), although with a slightly lower performance than the 10kb model, reflecting the challenges of annotating long-range regulatory elements. (**Figure 4a**). Taken together, we observed that Watershed-SV can prioritize rare functional SVs affecting either coding or non-coding sequence.

### Watershed-SV prioritizes structural variants implicated in Mendelian muscular disorders

To demonstrate Watershed-SV’s applicability to Mendelian disease diagnosis, we applied the method to a disease dataset with previously reported functional SVs implicated in patient phenotypes. Specifically, we applied Watershed-SV, trained using GTEx SRS SV calls and multi-tissue transcriptomic outliers, to prioritize functional rare SVs from 26 patients with inherited muscular disease, among which 2 are diagnosed with SVs and 6 with SNV from the Center for Mendelian Genomics (CMG) project [38]. We jointly called SVs using svtools [39] and Parliament2 [40]. After harmonizing variant calls with Jasmine, we called 11,360 deletions, 9,346 insertions, 3,820 inversions, and 592 duplications. Due to the challenges of calling rare SVs with a small cohort, we queried 1000 Genomes, CCDG, and gnomAD SV reference callsets with SVAFotate [41] and genotypes putative novel variants in 1000 Genomes SRS with PARAGRAPH [42] to annotate population SV allele frequencies (AF). Population database queries and PARAGRAPH enabled the annotation of AF in 97% of SVs. We called expression outliers in the CMG cohort using GTEx skeletal muscle as a reference expression baseline. We observed on average 318 outliers per individual in the CMG samples, in contrast to 94 outliers per GTEx healthy controls (**Figure S4a**). Using the Watershed-SV pipeline, we identified both cases where previously described diagnostic variants are SVs [38]; both are cases of Duchenne muscular dystrophy (DMD) caused by inversions in the DMD gene (**Figure S5e**). Notably, the patient C4 inversion included the first 18 exons, resulting in expression of a truncated transcript similar to other known cases of large-inversion induced DMD[43]; the C3 inversion inverted exon 51 and was associated with greatly reduced expression(**Figure S5e**).

Among 18 patients without prior genetic diagnosis, we observed four patients with Watershed-SV prioritized gene-SV pairs in muscular disorder related genes, one gene-SV posterior above 0.5, and three above 0.9. (**Table S6**). Among them, a female patient (N10) with undiagnosed Limb-Girdle Muscular Dystrophy carried a heterozygous rare deletion (gnomAD AF = 0.0003, Watershed-SV = 0.996, WGS-baseline = 0.78) disrupting the 3’ CDS-UTR region of *PIP5K1C*. Known disruptions of *PIP5K1C* can cause an autosomal recessive lethal contracture syndrome III (OMIM#611369) [44], another type of severe muscular disorder. While there is not sufficient evidence to determine that this deletion alone underlies the patient’s phenotype, it demonstrates Watershed-SV’s ability to prioritize variants of interest for further genetic and functional investigation.

### Watershed-SV prioritizes LRS SVs in rare disease

Knowing our method could prioritize diagnostic rare disease SVs from SRS, we investigate if we could prioritize likely functional LRS SVs that may underlie UDN patients’ disease phenotypes. We trained a 100kb Watershed-SV model that models per-tissue expression outliers from both blood and fibroblast using GTEx v8 data. We defined the Watershed-SV estimate as the maximum of the Watershed posterior probabilities for blood and fibroblast for each gene-SV pair. Out of 19,277 gene-SV pairs tested, 886 had a posterior of at least 60%, representing 4.6% of rare variants within 100kb of the nearest gene.

In order to assess the variants that Watershed-SV could return for clinical review, we evaluated variants passing a Watershed-SV score threshold along with filters corresponding to basic approaches clinicians could use to prioritize potential disease relevant variants. The filters encompass three general strategies: (1) filtering for variants near genes previously known to be related to the patients’ symptoms, using HPO term matching [45] (2) filtering for variants that have strong evidence of pathogenicity from WGS-based variant scoring, here using CADD-SV; and (3) prioritizing variants through RNA-seq outlier signals using expression outliers alone. Specifically, we compared Watershed-SV to CADD-SV [16], and evaluated whether Watershed-SV nominates different variants from CADD-SV, with these two scores evaluated alone and in combination with other filters. We compared a CADD-SV threshold of 10 to a Watershed posterior threshold of 0.6, corresponding to a roughly equivalent fraction (4.6%) of variants.

When comparing the most stringent set of filters, Watershed-SV combined with HPO term matching prioritized symptom relevant functional gene-SVs for more patients (n=6) than CADD-SV combined with expression outlier and HPO term matching (n=1) (**Fig. 5a**) – that is, among genes with established relationships to the patient symptoms, more hits were identified using Watershed-SV than among a comparable size set prioritized by CADD-SV. In addition, the variant gene-pairs prioritized by HPO term-matching along with either CADD-SV or Watershed-SV overlap minimally. When not constraining to expression outlier genes, filtering by HPO and Watershed-SV yields 18 variant-gene pairs, filtering by HPO and CADD-SV yields 12 variant-gene pairs, but the two sets only share a single variant-gene pair (**Figure S8a**). This suggests that Watershed-SV and CADD-SV contribute complementary information, with Watershed-SV optimized for prioritizing expression regulating variants and CADD-SV for constrained variants. Among all variants in UDN prioritized by either CADD-SV or Watershed-SV, we found an overwhelming majority coming from SVs and long indels (30-50bps) only identified in long read data (**Fig. 5b**).

**Figure 5.**
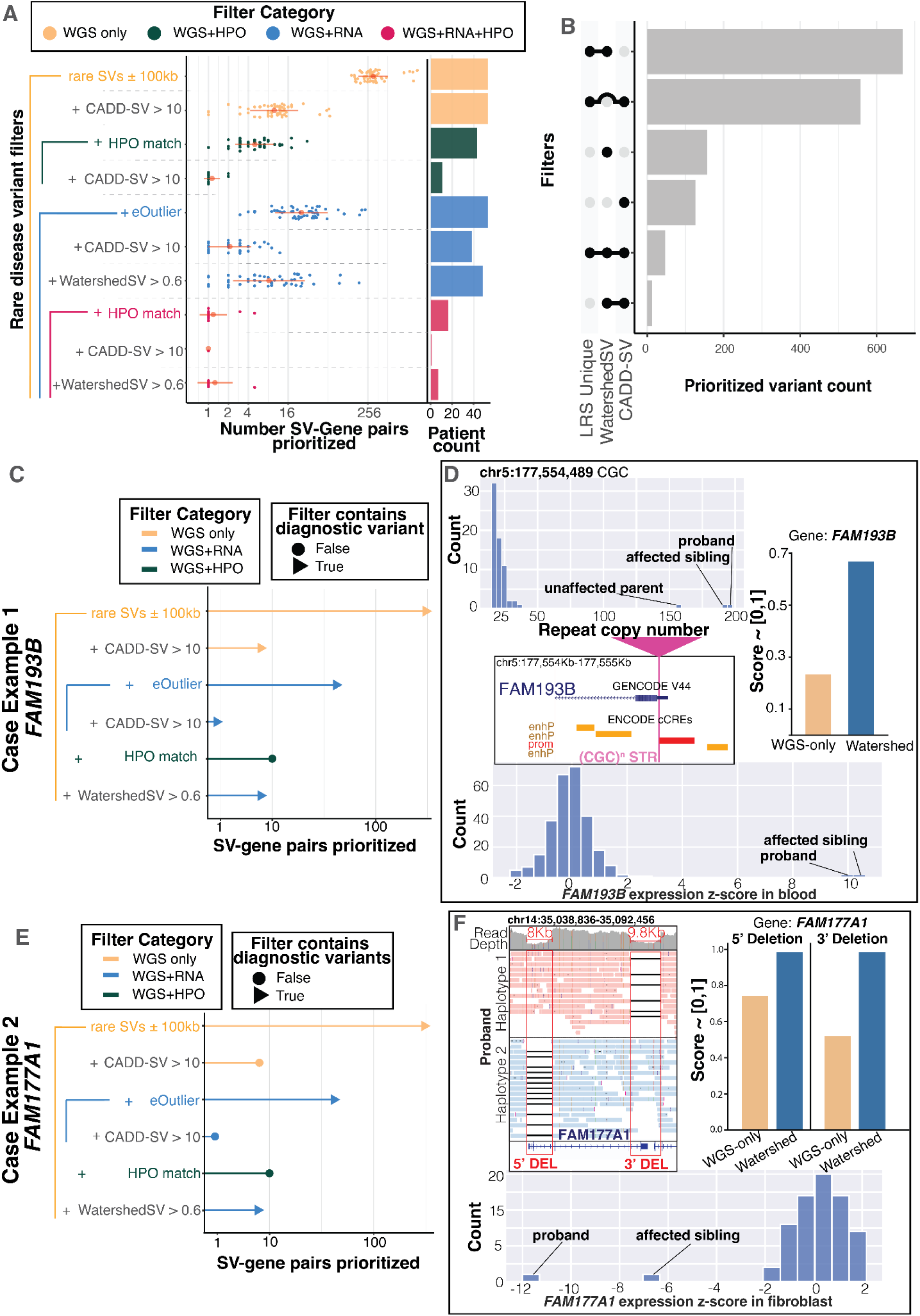
Watershed-SV prioritizes symptom-relevant functional rare SVs from UDN LRS dataset. **A** Swarmplot for number of gene-SV pairs prioritized per individuals in the UDN LRS dataset under different set of combined filters. There are 4 filter categories: WGS-only filters, WGS + HPO filters, WGS + RNA filters, and WGS + RNA + HPO filters, in increasing level of stringency due to increasing types of filters jointly applied; red dot represent the mean number of gene-SV pairs across individuals, red horizontal line represent standard deviation; x-axis is in log2 scale; the bar plot on the right shows number of samples with significant prioritizations. **B** Upset plot depicting number of gene-SV pairs prioritized by Watershed-SV (posterior > 0.6), CADD-SV (score > 10), and whether the SV is uniquely identified using LRS. **C and E** Case example 1, rare TREs shared by both siblings, and case example 2, rare compound heterozygous deletions in siblings. Lollipop plot shows which set of filter includes the candidate diagnostic gene-SV pair (Triangle) and which does not (Circle), height of the lollipop represents number of gene-SV pairs prioritized in log2 scale. **D** Panels depict the TR copy numbers of the siblings and unaffected parent with less-expanded allele. The TRE loci is in 5’ UTR of FAM193B. Both Watershed-SV and CADD-SV can prioritize this but not WGS-only model. Both siblings have extremely high overexpression z-scores. **F** Panels depict the compound heterozygous deletions phased onto both alleles for FAM177A1, causing LOF of gene and thereby underexpression outliers. Only Watershed-SV succeeded at prioritizing both variants.

Watershed-SV yielded a number of interesting variants for future analysis. In one case, we identified a heterozygous CCG TRE in a pair of siblings with clinically diagnosed oculopharyngodistal myopathy (OPDM). The TRE is situated in the promoter and 5’ UTR of the *FAM193B* gene and was observed with overexpression outliers in both of the siblings’ blood RNA samples. Previous studies have found CGG/CCG repeat expansions in the 5’ UTR of other genes to be causal for OPMD [46–48], and speculate repeat-associated non-ATG (RAN) translation of the CCG repeat and accumulation of toxic RAN proteins may be the mechanism. Fazal et al. predicted this repeat locus as pathogenic using RExPRT, a tool trained with tandem-repeat-specific annotations [49], but was unable to resolve the full size of the repeat using SRS. Comparing LRS vamos calls with ExpansionHunter calls at the same loci, we saw that short-reads drastically underestimated the length of the expansion (**Figure S9a**). Because *FAM193B* has not been annotated as a disease gene for OPDM, HPO-term-based filters failed to prioritize the variant. Both Watershed-SV (score = 0.63) and CADD-SV (score = 11) were able to prioritize the TREs (**Fig 5c**). We observed similar tandem repeat numbers (RN_A_=198, RN_B_= 194) for the two siblings and a smaller yet still expanded number in their unaffected parent (RN = 158), which could be consistent with a premutation allele that was expanded beyond a pathogenic threshold in both siblings (**Figure 5d**). CGG repeat expansions have previously been associated with hypermethylation and gene silencing, yet with nanopore methylation calls in this case, we did not detect a change in methylation levels of the *FAM193B* promoter, consistent with the observed over-expression(z-score = 10) (**Figure S9d**). Watershed-SV’s integration of gene expression as evidence of regulatory disruption provided greater support for the prioritization and pathogenicity of this variant.

In a second case, we detected a set of compound heterozygous deletions separately affecting the 5’ UTR-CDS region and the 3’ CDS-UTR region of *FAM177A1* in another pair of siblings suffering from global developmental delay, macrocephaly, and seizures. CADD-SV computed a modest score of just 6 for one of the variants, while Watershed-SV prioritized both variants with strong posterior probabilities > 0.9 (**Fig. 5e**). Notably, the siblings displayed strong under-expression in fibroblasts for FAM177A1 in comparison to other UDN samples. LRS enabled the phasing of these variants and showed the deletions were *in trans,* disrupting both *FAM177A1* alleles (**Fig. 5f**). A study [50] of disease variants in consanguineous families reported homozygous frameshift SNVs in *FAM177A1*(c.297_298insA) in individuals with similar neurologic symptoms to these siblings. Experimental validation study [51] using a Zebrafish model further supported that biallelic LOF mutations on *FAM177A1* could potentially lead to neurodevelopmental problems via abnormal Golgi complex dynamics.

## DISCUSSION

Multi-technology, high-sequencing-depth references have shown that there are roughly 22,000-27,000 rare SVs per genome [3,4]. We achieve similar numbers within our UDN samples using a multi-algorithm consensus set from only low-depth (15x coverage) long-read nanopore data, yielding on average more than 2.4x of what was found with SRS callers, especially substantially more insertions in LRS across all size ranges. Meanwhile, we observed more inversions, duplications, and large deletions >10kb in SRS than in LRS, likely due both to higher false positives in the SRS callset and limitations in alignment-based SV calling from LRS. From our study, we also demonstrated that LRS-based reference panels (ADRC) significantly improved variant allele frequency estimates, stressing the need for high-quality, diverse ancestry population reference such as the ongoing LRS sequencing of the 1000 Genome Consortium or All of Us [52,53]. In addition, we also examined variation at TR loci, genotyping TR copy number from LRS at 2 times more loci than profiled routinely in SRS, and identifying longer TREs even up to 10kb.

Beyond variant discovery, our work investigates the functional impact of LRS rare SVs and TREs with expression outliers measured from RNA-seq. We reinforce previous findings of a strong enrichment of gene expression outliers nearby rare SVs [9,13,14,54], and we further also detected expression effects of additional variant types, including insertions and TREs, and enrichment for non-coding rare SVs that disrupt important enhancers predicted by the ABC enhancer model [34]. Future investigation in larger cohorts with disease relevant transcriptomes and long-read genomes is expected to only further resolve the functional rare SV landscape. Relatedly, assembly-based SV callers like the Napu pipeline [55] and read-depth based SV callers may enhance future studies with increased sensitivity for detecting and interpreting other types of variation.

Finally, our work provides a framework, Watershed-SV (https://github.com/jasonbhn/Watershed-SV), to synthesize genomic annotations of SVs with transcriptomic signals to prioritize functional rare SVs, applicable to rare disease patients. We showed that Watershed-SV improves prioritization of rare SVs that affect gene expression over existing tools such as CADD-SV, and can identify candidate functional variants that would have been missed by such genome-only methods. Among the unique prioritizations from Watershed-SV in UDN LRS genomes, we successfully identified candidate diagnostic variants, suggesting the benefit of a dedicated model of rare SV’s impact on gene expression. However, current Watershed-SV models were trained on SRS data from GTEx, likely not capturing the full functional SV landscape observed in LRS. Future improvements may be possible from training on larger LRS datasets, and from incorporating additional molecular signals such as splicing, methylation available directly from nanopore sequencing, or proteomics [11]. Future improvements to Watershed-SV will also analyze transmission in trios or pedigrees of related individuals to further help variant prioritization.

In summary, we demonstrated the power of LRS in detecting functional and disease-relevant indels, TREs, and SVs using LRS genomes from UDN patients with negative ES or SRS. Altogether, our work is a demonstration of how LRS sequencing will broaden our understanding of the function of rare SVs and other rare variants in healthy and rare disease cohorts, and provided a reusable framework for practically integrating LRS SV with various transcriptomic outlier signals to provide additional candidate diagnostic variants for undiagnosed rare diseases. With the increasingly context-specific RNA-seq and the rapidly expanding LRS resources, our analysis framework will be an even more powerful tool for rare SV prioritization including for disease.

## METHODS

### Oxford Nanopore Sequencing

DNA was extracted from either whole blood (n=25) or from a cultured fibroblast pellet (n=43) for all UDN individuals using Qiagen’s extraction protocol (Cat. ID:158023). Up to 4µg of genomic DNA was used as input into Nanopore ligation kit LSK109 or LSK110. Standard ligation protocol was followed without barcoding. Libraries were loaded onto R9.4 flow cells on the PromethION device and sequenced to exhaustion. Libraries were washed and reloaded 24 hours into sequencing in order to maximize sequencing throughput. Some samples with lower depth were resequenced on additional flow cells to increase coverage. In total, 77 flow cells were used to sequence the 68 individuals. Data was basecalled in real time through MinKNOW with Guppy (v4.0.11), using the high-accuracy model (hac) with a Qscore filter of 7 for determining passed reads. Sequencing summary metrics were calculated and plotted with Nanoplot from sequencing summary files [56], and the region from GRCh38 aligned bam files to report percent identity.

### LRS Structural Variant Calling

FASTQ files from base calling were aligned to the GRCh38 patch 13 reference (https://www.ncbi.nlm.nih.gov/datasets/genome/GCF_000001405.39) with minimap2 (2.26-r1175) using the –map-ont presets after filtering low quality (Qscore < 7) and short-length reads (length < 250bp) [57,58]: `“zcat $fastqs | NanoFilt -q 7 -l 250 | minimap2 -x map-ont -a --MD -t {threads} {input.ref} │ samtools view -hb > {output.bam}”`. Structural variants were called with an ensemble algorithm combining three separate SV callers: Sniffles2 (v2.2) [22], cuteSV (v2.1.0) [23], and SVIM (v2.0.0) [24]. Each tool was called with the following parameters:

**Sniffles**: `“sniffles -t {threads} 4 --sample-id "{sample}_sniffles"`

`--input {input.bam} --minsvlen 30 --minsupport 3 --vcf {output.vcf}`

`--output-rnames --tandem-repeats {ttrf_bed}”`

**CuteSV**: `“cuteSV -t 4 --sample {wildcards.sample}_cuteSV`

`--max_cluster_bias_INS 100 --diff_ratio_merging_INS 0.3`

`--max_cluster_bias_DEL 100 --diff_ratio_merging_DEL 0.3`

`--report_readid --min_support 3 --min_size 30 --max_size 10000000`

`--genotype {input.bam} {input.ref} {output.vcf} {params.scratchdir}”`

**SVIM**: `“svim alignment --sample {sample}_SVIM --minimum_depth 3`

`--min_sv_size 30 --max_sv_size 10000000 --read_names {params.tmpdir}`

`{input.bam} {input.ref}”`

Additionally, for SVIM output vcf files were processed to ensure consistent naming of duplications across tools and consistent names for reporting supporting read IDs to be used downstream for Iris insertion refining. After calling SVs with each tool, individual callers per sample were merged with Jasmine SV (v1.1.5)[25]. For individual merges we used the following Jasmine parameters: “jasmine -Xmx18g --allow_intrasample --normalize_type --ignore_strand --file_list={params.vcf_list} --output_genotypes --centroid_merging --dup_to_ins min_support=2 genome_file={input.ref} out_dir={params.tmp_dir} out_file={out.vcf}”.

Genotypes were decided for the post-merged consensus by taking a majority vote of the genotypes determined by each of the three callers. We required that a structural variant be supported by at least two callers, creating the consensus set we used for downstream analysis. After merging with Jasmine, we also ran Iris through Jasmine in order to refine insertion sequences of noisy Nanopore data. We ran Iris refinement with these parameters: “jasmine -Xmx 24 --preprocess_only --run_iris iris_args=keep_long_variants, vcf_out={output} genome_file={input.ref} file_list={params.vcf_list} bam_list={input.bam_list} out_dir={params.tmp_dir} out_file={output.vcf} threads=4”.

### SRS Structural Variant Calling

For short-read sequencing (SRS) structural variant calling, we sought to emulate what is typically performed for clinical WGS genetic testing. The current clinical standard for WGS clinical testing uses the Illumina DRAGEN pipeline [59] and calls structural variants with Illumina’s Manta software. We ran Manta (v1.6.0-2) with default parameters. To limit false positive calls, which are prevalent in short-read call sets, we ran an additional genotyping step with PARAGRAPH. First, individual Manta calls across the UDN cohort were merged with JasmineSV using the following parameters: `“jasmine file_list=∼{UDN_manta_file_list} out_file=∼{UDN_SRS_cohort_vcf} –output_genotypes –default_zero_genotype”`. Using this merged set, we then input fully resolved SVs into a PARAGRAPH genotyping step. Insertions are not fully resolved above twice the fragment size of the library according to Manta documentation (<u>github.com/Illumina/manta/blob/master/docs/userGuide/README.md#known-limitations</u>), thus these unresolved SVs could not be genotyped with PARAGRAPH. We ran PARAGRAPH on each SRS BAM file with the following parameters: “*`idxdepth -b ∼{cram} -r`* `∼{reference} -o ∼{sample_id}.json`”. To estimate the average depth across the genome, we used the following parameters: “*`multigrmpy.py -i ∼{vcf} -m`* `∼{sample_id}.manifest.txt -r ∼{reference} -o ∼{sample_id} -M 20*{depth}`”. -m is the manifest file extracted and formatted from idxdepth output, and -M, maximum allowed read count is set according to average depth across the genome. WDL workflow is available on Firecloud and AnVIL [60] (https://portal.firecloud.org/?return=anvil#methods/run_paragraph/run_paragraph/26). From this final merged, genotyped callset, we calculated variant lengths and counts per sample across structural variant types for 50 UDN individuals that had both WGS short-read and long-read data available.

### Allele Frequency Estimation

To assess structural variant allele frequency we first applied SVAFotate [41]. SVAFotate uses a large population reference of SVs from SRS that includes gnomAD, CCDG, and 1000 Genomes, and matches an input SV to the catalog given specific overlap fractions to assign input SVs with allele frequencies from the SV catalog. We ran SVAFotate on the UDN short-read Manta-Jasmine-merged callset, UDN long-read-consensus callset, and IMD short-read SVTools-Parliament2-Jasmine-merged callset with the following parameters: “*`--a best –f 0.5`*“ . For the short-read callset, we saw high ascertainment of variants in the SVAFotate catalog, 88%. We also ran SVAFotate with the same parameters on the Jasmine-merged consensus set from Nanopore sequencing; however, observed drastically lower ascertainment of variants, 8%.

To further assess variants not detected with SVAFotate, we genotyped all no-hit variants with PARAGRAPH genotyper in 2,499 high-depth 1000 Genome unrelated individuals, which allowed us to obtain allele frequency estimates for 97% of SRS variants and 92% of LRS variants. To get more robust allele frequencies for ONT and address variants outside of the short read detection limit using PARAGRAPH, we used consensus SV calls from long read sequencing of Stanford’s Alzheimer’s Disease Research Center (ADRC) and the Stanford aging and memory study (SAMS) (n=571) sequencing collection[32]. The ADRC contains 598 individuals sequenced on the same Oxford Nanopore PromethION platform to a median of 60 Gb of coverage. An identical SV calling and merging workflow was run on ADRC. To get allele frequencies we ran a population Jasmine merge on all genomes from the ADRC and UDN (667 total genomes) using the following jasmine parameters: “*jasmine -Xmx96g -Xms24g --ignore_strand --file_list={params.vcf_list} --normalize_type --dup_to_ins --default_zero_genotype --centroid_merging out_dir={params.tmp_dir} genome_file={input.ref} --output_genotypes out_file={output.vcf} threads={threads}*”. Merging was performed per chromosome to decrease memory requirements. From the population-merged vcf, allele frequencies were estimated by summing allele counts identified in the merged vcf and dividing by total allele number.

### Tandem Repeat Copy Number Genotyping

For long-read sequencing data, the vamos efficient motif set was downloaded from (https://github.com/ChaissonLab/vamos/blob/master/snakefile/configs/vntr_region_motifs.e.bed.gz), and used as input into vamos (v1.3.6) to genotype tandem repeat (TR) copy numbers across across 467,000 TR loci [27]. vamos was called on each sample using the following parameters “*`vamos --read -t 4 -b {input.bam} -r vntr_region_motifs.e.bed -s {sample_name} -a 0.93 -o {output.vcf}`*”. Vamos vcf files were converted to tsv files to report per sample and haplotype length of each TR (vcf INFO FIELD LEN_H1, or LEN_H2). We considered only repeat annotation lengths and not composition information. TRs were annotated by motif size based on the length of the first motif present in the efficient motif catalog.

For short-read sequencing data, TR copy numbers were estimated with ExpansionHunter (v5.0.0) [28]. The recommended TR catalog for ExpansionHunter was downloaded from (https://github.com/Illumina/RepeatCatalogs/blob/master/hg38/variant_catalog.json). ExpansionHunter was ran with the following parameters: “*`ExpansionHunter –reads {input.bam} --threads 4 --reference {input.fasta} --variant-catalog variant_catalog.json --output-prefix {output_prefix} --analysis-mode streaming`*”. The resulting output json was converted into a tsv file to report per sample per haplotype estimated repeat numbers and confidence intervals. TRs were again annotated based on their motif length.

For both short reads and long read TR files, TR tsv files were concatenated across all samples within each technology, we then grouped by unique TR loci and calculated mean and variance per TR across all alleles. TRs were filtered to those that were detected in at least 50% of UDN individuals and detected in at least 50 haplotypes. This resulted in 466,220 TR loci for vamos and 174,260 for ExpansionHunter. For comparing the ExpansionHunter variant catalog to the vamos TR catalog, we calculated genomic ranges based on coordinates found in their respective catalogs, and we flanked each region by 100bp on each side. The ExpansionHunter and vamos motif sets were then intersected with bedTools to determine if they were covering the same loci. For loci in both catalogs that did intersect, we further characterized if they contained the same motif. Vamos includes potentially multiple motifs at each TR in order to represent composition changes that occur within repeats, whereas ExpansionHunter only records a single motif. To compare if ExpansionHunter and vamos contain the same motif, we compared the ExpansionHunter motif with any motif in the vamos set, or any cyclic permutation of a motif in the vamos set (because, for instance, a CGC repeat and a CCG repeat would result in identical repeat sequence).

To characterize tandem repeat variation at known pathogenic loci, we downloaded the STRchive motif set from (<u>github.com/hdashnow/STRchive/blob/main/data/hg38.STRchive-disease-loci.TRGT.bed</u>). And reformatted this file so it is compatible to be run with vamos and ExpansionHunter. We analyzed the pathogenic motif set with ExpansionHunter and Vamos as described above. The resulting haplotype-level tsv files were concatenated across the cohort in each technology and then joined on TR_id. Since assignment of haplotypes in both tools is arbitrary, to compare haplotype-specific repeat copy numbers, we assign H1 to be the haplotype with the longer repeat count (max) and H2 to be the haplotype with the shortest repeat count (min). TRs were filtered if there was >50% missingness in individuals, removing 20 TRs from ExpansionHunter. For the remaining TRs, EH mean and vamos mean repeat copy numbers were calculated, and per loci Spearman correlations between reassigned (max,min) haplotype repeat copy numbers were computed.

### Mean Neighbor Distance Rare Tandem Repeat Expansion Calling

To detect rare and extreme tandem repeat expansions (TRE) at tandem repeat loci, we developed a method which involves calculating and thresholding on a mean neighbor distance (MND) statistic. This semiparametric approach uses a population reference (preferably of healthy controls) to jointly define robust tandem repeat copy number distributions together with any samples of interest. Then for each observation in the distribution (corresponding to a unique TR copy number allele observed in one haplotype of an individual) we calculate how far on average they are from their K-nearest neighbors (mean of the minimum K absolute differences). We then standardized this mean neighbor distribution by the standard deviation of the original distribution. This statistic describes how many standard deviations away an allele is from their nearest K-neighbors. Here, if an allele was drawn from the same distribution as the rest of the alleles, then its nearest neighbors would be very close, and the standardized MND would be close to zero. Even if an individual was at the tail of the distribution, there would still be neighbors nearby and the standardized MND will still be close to zero. However, if an allele was extremely expanded, as is the case in repeat expansion disease, they will be sampled from a distinct distribution and should be clearly separated from the rest of the distribution. This means the k-nearest neighbors should be far away and the MND will be high. Thus, thresholding on a standardized MND can differentiate extreme expansions from just individuals who are near the tail of the distribution (see schematic in **Figure 1B**).

This MND method is analogous to a z-score, but instead of calculating difference from an observation to the population mean, we replace the population mean with a local mean of the k-nearest neighbors of the observation. Thus, MND can be thought of as a topological or local z-score. Additionally, to require the repeat expansion to be at the tail of the distribution and not at an intermediate value between two mixtures, for instance, we also require the observation to be in the top K in order to be considered an expansion outlier (or bottom K in the case of rare extreme contractions). Finally, to avoid inflation of standardized MNDs from TRs with little variability, we forced a lower bound on the standard deviation of the original distribution by the expected standard deviation if the distribution was Poisson distributed, namely the standard deviation of the mean.

We observed that this method works well for detecting rare and extreme repeat expansions. Benefits of this model included its nonparametric estimation of the MND based only on nearest neighbors. This makes our method less susceptible to violations of underlying assumptions like the unimodality of data that is often assumed for standard z-score scaling outlier methods, an assumption commonly broken by the multimodal distributions of many TRs. Also outlier calling methods like Tukey’s assume a wide variance of the distribution in order to calculate interquartile ranges, however, many tandem repeat loci are pathological in this regard as the 25th and 75th percentile will have the same value, while still being variable in the remaining 25% of data points, thereby making any other value an outlier.

To call repeat expansions with this method there are two parameters to set. First, K to determine the number of nearest neighbors to consider when calculating the local z-scores. As K increases, the local z-score will approach the standard global z-score and local information about the distribution will diminish. A default k parameter propotional to the total allele number in the reference population is recommended, for instance K = 0.5% of total alleles. This mirrors rare variants at a < 0.01 MAF, where if we see 1% or more of the alleles having the same or similar repeat number to a given allele, it will have a MND close to zero and will not be called a TRE. For determining ultra-rare events, a lower k threshold of 0.5% or 0.01% of total allele number can be used. Whereas more lenient tandem repeat expansion calling can be done at 2%. In the most extreme case, if K = 1, then an allele is only an outlier if no other allele is nearby, which should only be done in a rare disease setting if it is absolutely sure no other allele could have the same pathogenic repeat. We show increasing K results in more TREs called on average per sample (**Supplemental Figure 3A**). The second parameter is the standardized MND threshold above which to call a sample an rare TRE. We used a default parameter of 2, but this can be increased in order to be more stringent and detect only the most extremely expanded of alleles. We also show that increasing this standardized MND threshold results in less and less TREs called per genome on average (**Supplemental Figure 3A**).

### Rare tandem repeat expansion calling in UDN

We applied the above mean neighbor distance (MND) method to vamos (from LRS genomes) and ExpansionHunter (from SRS genomes) tandem repeat genotypes from the UDN to call extreme tandem repeat expansions. In the first round of analysis we calculated the MND on the default catalogs present for each tool. For the LRS vamos set, we jointly analyzed the data with ADRC vamos genotypes as a reference set, whereas for the SRS ExpansionHunter, we used the ExpansionHunter repeat catalog that was run on 1000 Genomes as a reference set, downloaded here (<u>github.com/Illumina/RepeatCatalogs/blob/master/hg38/genotype/1000genomes/1kg.gt.hist.tsv.g</u> <u>z</u>). To call repeat expansion outliers, we set K = 0.5% of total allele number, corresponding to K = 5 in the vamos set, and K = 25 in the ExpansionHunter set. We set a standardized MDN threshold of 2 above which to call an allele as an extreme tandem repeat expansion (TRE). In a secondary round of analysis, we called TREs specifically from the STRchive catalog of pathogenic repeats. We used the same vamos and ExpansionHunter output as described above to input into the MND TRE calling script. For ExpansionHunter, because the 1000 Genomes reference repeat catalog did not contain all of these pathogenic repeats, we used a reference set of an additional 54 short-read UDN genomes that were also called with ExpansionHunter (bringing total reference size for SRS to 104). To account for the smaller reference size of ExpansionHunter, we decreased k for calling from the pathogenic repeats to k = 5 for the ExpansionHunter set (∼2% of the total allele number).

In order to compare our MND statistic to other outlier calling methods for tandem repeat expansion calling, we also called expansions from vamos output using two additional methods. We used the joint distribution of UDN combined with ADRC to perform all tandem repeat expansion calling. A Tukey’s outlier method was applied by setting an outlier threshold per TR equal to the 75th percentile of the distribution + 1.5*IQR of the distribution. Any allele that had a repeat count greater than this value was called an outlier. We also used a z-score scaling method where repeat counts per tandem repeat were scaled to a mean of 0 and standard deviation of 1 by subtracting the population mean and dividing by the population standard deviation. A z-score threshold of 3 was used to compare with Tukey’s and MND threshold. We compared counts of tandem repeat expansions detected by each method from the UDN genomes and found MND drastically decreases the number of TREs compared to Tukey’s method. Compared to the z-score method, the MND method calls about half as many TREs as z-scores (**Supplemental Figure 3C**).

### Outlier calling against a healthy population background in matching tissues

In the rare disease datasets we collected 24 skeletal muscle biopsy RNA-seq STAR-mapped bam files from the CMG Muscular Disease dataset [38] and 66 fibroblast and 283 blood samples bam files from the UDN dataset. In order to more effectively identify aberrant expression from healthy samples, we also selected GTEx RNA samples from skeletal muscle, cell cultured fibroblast, and whole blood as the best matching tissues controls. To reduce technical variation due to sample quality issues, we filtered all RNA-seq based on RNA Integrity Number (RIN) greater than 5.5, To quantify expression on a gene level, we used RNA-SeQC to generate read count and TPM [61] with GENCODE[62] models matching the versions the STAR alignment was performed on. We limited, for a given tissue type, the genes to those with at least 6 reads in at least 20% samples. In UDN blood samples, to maximize discovery power, GLOBINclear was used on cDNA library to maximize the number of non-globin and disease relevant mRNA to be detected. This created a library compositional shift when comparing GTEx whole blood and UDN globin-depleted blood. To combat the composition bias, we generated TMM adjusted TPM values using edgeR package, accounting for this known unwanted shift in RNA composition that TPM normalization itself cannot account for [63]. After acquiring TPM values, we estimated expression PCs, which was previously shown [64] to well-represent the technical hidden covariates. TMM-TPM are log transformed using log_2_(TPM+2), then scaled to mean of 0 and standard deviation of 1. In addition to 60 PCs [9], we also accounted for batch, RIN, and biological sex using linear model, and subsequently scaled the expression residual from the model to obtain expression z-scores. We removed global outliers in similar fashion as Ferraro et al. 2020 [9].

### Expression outlier enrichment nearby rare SVs analysis

In order to test enrichment of expression outliers nearby rare SVs we used a multivariate logistic regression framework, using outlier status as the dependent variable and presence of rare SV covering various annotations nearby the gene as independent variables. We performed enrichment in UDN using blood expression z-scores as calculated above to maximize sample size and power. After filtering out individuals who were global expression outliers, defined as individuals with number of outliers greater than 1.5*interquartile range + 75 percentile of outlier counts in the cohort, there were 33 UDN individuals with LRS and blood z-scores. We set a window size of 10kb to determine if an SV is near a gene, and intersect SVs with these expanded gene windows using bedtools[65]. To set the proper background, we restricted to genes with at least one outlier among the 33 individuals. In order to test SV Type enrichments, we set various increasing absolute z-score thresholds to determine a binary gene outlier variable, and then ran the following logistic regression to estimate log odds-ratios **Outlier ∼ SV_TYPE_DEL + SV_TYPE_INS + SV_TYPE_DUP + SV_TYPE_INV.** Where each of the dependent variables is set to 1 if the gene has a nearby SV of that type within 10 kb and 0 otherwise. For testing rare TREs we run a similar univariate model **Outlier ∼ has_TRE,** where has_TRE is set to 1 if there is a rare TRE present within 10kb of the gene. We also run specific models stratifying by Outlier direction, **Underexpression_Outlier** ∼ **SV_TYPE_DEL + SV_TYPE_INS + SV_TYPE_DUP + SV_TYPE_INV,** where Underexpression_Outlier is set to 1 if z-score is strictly less than negative of the Z-threshold used. And **Overexpression_outlier ∼ SV_TYPE_DEL + SV_TYPE_INS + SV_TYPE_DUP + SV_TYPE_INV,** where Overexpresion_Outlier is set to 1 if z-score is strictly above the Z-threshold used. The logistic regression estimates a beta value, which corresponds to the log odds ratio of that variant type being enriched for expression outliers compared to inliers, jointly controlling for the effects of the other variant types. We also extracted standard errors of this beta value and a p-value for significance. P-values were then Bonferonni adjusted to account for multiple hypothesis testing correction.

For testing enrichments across rare variant annotations, we used an absolute z-score threshold of z = 4 and increased window size to 100kb for determining SVs proximal to genes so we could capture more noncoding variants. Annotations for enrichment analysis were consistent with those that were used as features for Watershed-SV (details below). To create ABC annotations, we downloaded cell-type specific ABC elements from (ftp.broadinstitute.org/outgoing/lincRNA/ABC/AllPredictions.AvgHiC.ABC0.015.minus150.ForABCPaperV3.txt.gz), and cell-types were subset to those from primary, unstimulated cell types from blood, fibroblast, or muscle cell types in order to match the expression tissues we profiled in this study [34]. Coordinates for ABC elements across all tissues were then merged after grouping by ABC element class (promoter, intergenic, genic) and target gene and then using plyranges[66] *reduce_ranges* to create a tissue-level reference. The target gene column of ABC elements was used to map the ABC element to the gene it is predicted to be regulating. These coordinates were then lifted-over to hg38 using “*`CrossMap region ∼{chain_file} ∼{ABC_enhancer_bed} ∼{ABC_enhancer_lifted_bed} -r 0.85`* ” to be in a consistent build with SV coordinates.

For VEP gene body region annotations, VEP categories were made exclusive using the following prioritization schema [in order of highest priority: exon variant, 5’ UTR variant, 3’ UTR variant, intron variant, upstream noncoding variant, downstream noncoding variant]. So any SV that overlapped multiple features is assigned to the category with highest priority, making gene body region categories not correlated. Gene body region annotation enrichments were calculated by the following model: **Outlier ∼ upstream_noncoding_variant + five_prime_UTR_variant + exon_variant + intron_variant + three_prime_UTR_variant + downstream_noncoding_variant** in order to jointly estimate the enrichment of each gene body region. For length enrichments, we label genes as 1 if it has a rare SV above the given length threshold in a 100kb window and 0 otherwise, and compute enrichments from the model **Outlier ∼ has_SV_above_length,** using length threshold set to 100bp,1kb,10kb, and 100kb. CADD-SV [16] scores were generated for insertions, deletions, and duplications (CADD-SV could not score inversions), using their recommended snakemake pipeline and using as input the UDN plus ADRC jasmineSV-merged combined vcf [16]. Similar enrichments to SV length were performed using the CADD-SV score as a threshold and the model **Outlier ∼ has_SV_above_CADDSV** using the following thresholds of CADD-SV [5,10,15,20]. Similarly for VEP impact categories, enrichments were calculated with the model **Outlier ∼ has_MODIFIER_SV + has_MODERATE_SV + has_HIGH_SV.**

Finally, to look at regulatory annotations, we compiled noncoding regulatory annotations as described above for ABC elements and below for the other Watershed-SV features. SVs that intersected ABC elements of the target gene were assigned a 1. For non-binary features, i.e. LINSIGHT, PhastCon, REMAP CRM density, high and low categories were assigned based on whether the non-coding variant was above the median value of these annotation categories across all SVs with annotation present. We set the regulatory annotation feature to 0 if the SV also overlaps an exon, so they are exclusive to coding variants and to prevent them from spuriously capturing coding variant effects. We also created an annotation of putative-regulatory SVs for SVs that overlap any non-coding annotation or the VEP regulatory region variant annotation. We also create putatively non-regulatory variants as a control, which are variants that do not overlap any of the regulatory annotations and additionally do not overlap any high CADD-scoring variants, and the mean CADD score of the highest-CADD-scoring 10 SNVs overlapping the SV was less than 5. Individual SV annotations were then merged on a per-gene-individual level to get gene-level annotations. Due to high correlation among the different regulatory annotations, we tested enrichment for each annotation individually rather than jointly modeling them because beta values of correlated features in multivariate regressions are not interpretable. So, for each regulatory annotation, we compute enrichments by the following model: **Outlier ∼ has_coding_SV + has_SV_with_regulatory_annotation,** controlling for if the gene has a coding variant to avoid non-coding enrichment just from tagging coding SVs that are on same haplotypes. We report the beta value for the **has_SV_with_regulatory_annotation** term as the log odds ratio enrichment. Again, as with the SV Type models, we used the beta estimate as well as the standard error and p-value to report enrichments. We adjusted p-values using the Bonferroni method to correct for multiple hypothesis testing.

### Watershed-SV annotation collection pipeline

Given the increased complexity of annotating SVs relative to SNVs, Watershed-SV includes a more comprehensive set of relevant annotations: VEP [33] variant category and protein-coding sequence consequence, SV specific annotations like variant types and length, chromHMM [67] states, and conservation scores, among others. Since many SVs affect a range within the genome, leading to overlaps with multiple annotations from the same source, we summarized the impact of these SVs on nearby genes that aligns with increased functional impact for a given annotation. Since there could be more than one rare SV nearby a given gene, we further aggregated annotations according to **Table S5**, highlighting the strongest impact of variants nearby gene. Both gene level predictions and gene-variant level predictions can be computed for an individual from the trained model.

Further, two annotation collection methods are provided. First, we have a mode that adds user specified flanking sequence to each end of a given gene, then considers the aggregated impact of all rare SVs overlapping these regions as the genomic annotation. In this version, variant in gene body, upstream flanking region (UFR), and downstream flanking region (DFR) are aggregated together, regardless of the specific location of variant overlap. In another mode, we separately consider nearby the gene body, UFR, and DFR regions in order to better isolate the impact of rare SVs on different regions that might be contributing to outlier gene expression. In both modes, annotations are only collected from the overlapping region between an SV and a gene with its +/- flanking sequences to limit the impact to cis-regulatory. Our pipeline is available at (https://github.com/jasonbhn/Watershed-SV).

### Watershed-SV annotation features

#### SV-generated features

We one-hot encoded the type of rare SVs present nearby a gene like a presence-abscence vector. Allele frequency, length of the SV, copy numbers of variants could both be processed separately and uploaded as TSV or extracted from VCF file automatically. We log transformed length since it’s a continuous positive feature in our feature encoding. AFractions of exons sequence affected by nearby SVs were calculated and aggregated using the maximum among SVs to depict the most deleterious impact. Similarly, binary features describing exon truncations by SV from either 5’ or 3’ of the genes were included. For non-coding variants, distances of variants to UFR and DFR were calculated, with the aggregated minimum among nearby SVs being used for gene-level feature curations; note, when variant is coding, these distances are 0.

#### VEP features

We one-hot encoded the Consequences from VEP into binary features. We replaced the exon_variant, intron_variant consequences from VEP with our own script because the VEP module can simultaneously call an SV being intronic and exonic, producing contradictory information for model training.

#### Regulatory element annotations

In order to capture the impact of rare SVs on various regulatory elements, we used annotation from multiple sources. Activity-by-contact(ABC) model predictions were extracted and aggregated from 78 cell lines and tissues, we considered any rare SVs nearby genes that overlap an ABC enhancer mapped to the given gene an ABC SV. We extracted cis-regulatory modules(CRMs) from REMAP2022 database [35] and considered the maximum SV disrupted CRM score the remap score annotation for a gene. And we also collected information about tissue specificity of enhancers in the form of the number of primary tissues a given enhancer bed segment is detected in from enhancerAtlas 2.0 [68]. Finally, the number of tissues a TAD boundary is detected in Wang et al [69] are collected to depict the impact of SV on regulations related to 3D genome organizations.

#### ChromHMM features

To collect chromHMM features, we selected 27 primary tissue/cell types from 127 epigenomes. We used the 25 state model generated by the Roadmap Epigenome consortium[70]. We aggregated overlapping segments of the same state from 27 tissues into a single segment, with the count of tissues that the state is active in, forming modules of chromatin states. The higher the number of tissues, the more ubiquitously observed a state is.

#### Conservation Scores and other bigwig track features from UCSC Genome Browser

We used pybigwig[71] to collect and summarize the score tracks of various conservation metrics, including LINSIGHT, CADD, PhastCON, GC-content, and percent CpG in a disrupted CpG island from the UCSC Genome Browser[72]. LINSIGHT, CADD, PhastCON have high sparsity in the scores, therefore, we selected the top 10 scores within the range of each SV, and took the average of the top 10 as the annotation for these score tracks. For GC-content, the mean of GC was taken. And for CpG, the max CpG percentage was considered.

### Region-specific annotation aggregation

For region-specific annotation, conservation scores, regulatory annotations, chromHMM features are summarized and aggregated in the gene body, UFR, and DFR regions separately. Variant type information is also collected such that if an SV overlaps with the gene body and UFR, the SV segments that overlaps each regions will be considered separately, i.e. SVTYPE: DUP_UFR, DUP_gene_body. Other annotations, such as VEP, are aggregated as we did in non-region-specific aggregation.

Methods, scripts, and source annotations are available on github (https://github.com/jasonbhn/Watershed-SV). All scripts for each type of annotations could be ran independently, although we provide bash scripts and snakemake pipeline template to easily run on different VCF and outlier data types.

### Evaluating Watershed-SV using N2-pairs in GTEx samples

For evaluating the Watershed model, we largely followed the N2-pair evaluation framework developed previously [9]. To further account for variability within a single locus, specifically CNVs with multiple possible rare copy numbers, we required that copy numbers must at least match in the direction of copy number change from the population mode copy number. To account for noise in the gene expression outlier z-scores, we evaluated using stringent outlier signals threshold at Z>3, but relaxed the outlier threshold to a p-value threshold of 0.05 in the second individual in N2 pairs. To evaluate the performances of models on non-coding variants, we performed the same analysis but limited to only N2-pairs with non-coding rare SVs nearby. We further relaxed the p-value threshold to 0.1 due to the weakened impact of non-coding variants in comparison to coding variants (**Supp. Fig. 5c**). Bootstrapping of AUPRC is performed identically as mentioned in previous literature.

### Applying Watershed-SV models to prioritize functional rare SVs in the inherited muscular disease dataset

Structural variants were called using svtools[39] and Parliament2[40]. svtools jointly infers the presence of SVs based on LUMPY[73] breakpoints and CNVnator[74] copy number changes. Due to the small cohort size, this approach is underpowered in calling rare duplications or deletions. We used Parliament2 on each sample, which merges variant calls from multiple callers for a single individual, to maximize recall of rare and private SVs that failed to be called from SVtools. Due to the high false positive rate of SV calling in short read data, we required an SV to have the support from at least two different callers [40]. Subsequently, we merged individual sample VCFs using Jasmine. Finally we harmonized the Parliament2 and SVtools callset using Jasmine. Variant allele frequencies were ascertained using SVAFotate and PARAGRAPH as described earlier. We collected annotations for rare SVs found in the dataset, combined with skeletal muscle expression outliers to run Watershed-SV prediction mode based on the GTEx 100kb region specific median z-score model.

### Applying Watershed-SV models to prioritize rare SVs and extreme TR expansions in UDN dataset

We applied both the median z-score model and the tissue-Watershed model that was trained jointly with blood and fibroblast outlier signals to the patient data. For rare SVs, predictions were performed as described in the previous section. For extreme TR expansions, we cast them to the type of duplication, and calculated the length as the total length of the expanded repeat. To evaluate whether Watershed-SV identified symptom relevant rare disease variant gene pairs, as well as to evaluate performance against CADD-SV, we designed a set of filters to simulate a range of diagnostic filters that genetic counselors would use. The filters encompass three general approaches: matching of nearby gene with the patient’s symptoms, variants that have strong evidence of pathogenicity from whole genome sequencing based variant prioritization methods, and variant prioritization leveraging RNA outlier signals. We compared CADD-SV to Watershed-SV to assess how CADD-SV scores for the same set of variants nearby 100kb of genes with measured expression in blood or fibroblast compared to Watershed-SV scores. We note that CADD-SV is a variant centric prioritization approach and does not model the variant impact on a nearby gene. Furthermore, the CADD-SV score is on a Phred Scale whereas Watershed-SV reports a probability. The same CADD-SV score is used for two gene-SV pairs that are the same SV but different gene in Watershed-SV. We used a Watershed threshold of 0.6, corresponding to ∼10% of the total variants, and a roughly equivalent threshold of 10 in CADD-SV, corresponding to the top 10% pathogenicity. To compare whether variants prioritized are relevant to disease phenotypes, we created HPO term matching using Phen2Gene [45], considering all genes for each patient with rank < 300 as recommended by authors.

## Data Availability

All UDN data produced in the present study are being uploaded to dbGaP under the accession phs001232. All GTEx data are under dbGaP accession phs000424.

## DATA ACCESS

Expression outlier z-scores from GTEx were deposited in dbGaP (phs000424). Outlier z-scores, harmonized variant VCFs with genotype information for UDN data were deposited in dbGaP (phs001232), along with bam files from ONT long read sequence alignment. Prioritized top hits with no personal information are released in the supplementary table S7.

## ACKNOWLEDGMENTS

We would like to thank Benjamin Strober, Taibo Li for suggestions on Watershed-SV model, Joshua Weinstock and Rebecca Keener for editing of this manuscript and helpful conversations related to this work. Research reported in this manuscript was in part supported through the Undiagnosed Diseases Network (Award U01HG010218) and the GREGoR Consortium (Award U01HG011762). TDJ is supported by U01HG011762, T32HG000044. BN is supported by R35GM139580, U24HG010263, OT2OD034190, U01CA253481, R03CA272952, U01HG012069. JEG is supported by U01HG010218, U01HG011762. SF is supported by 1R21HG013397, 5R01NS072248. CMR is supported by U01HG010218, U01HG011762. DEB is supported by U01HG011762, U01NS134358. RAU is supported by U01HG011762. PCG is supported by U01HG011762. ANR is supported by U01HG010218. EAA is supported by U01HG010218, U01HG011762 JAB is supported by U01HG011762 and U01NS134358. SZ is supported by 1R21HG013397, 5R01NS072248. MDG is supported by R35AG072290, P30AG066515, R01AG074339, R01AG048076. SBM is supported by U01HG011762, U01AG072573, R01AG066490, R01MH125244, and U01HG012069. MCS is supported by U24HG010263, OT2OD034190, U01CA253481, R03CA272952. MW is supported by U01HG010218, U01HG011762. AB is supported by R35GM139580, U01HG012069. This work utilized computing resources provided by the Stanford Genetics Bioinformatics Service Center, supported by NIH Instrumentation Grant S10OD025082, and would not have been possible without the support of the Stanford SCG cluster system administrators.

