## Supplementary Figures for "Integration of transcriptomics and long-read genomics prioritizes structural variants in rare disease"

### UDN Watershed Supplementary Figures

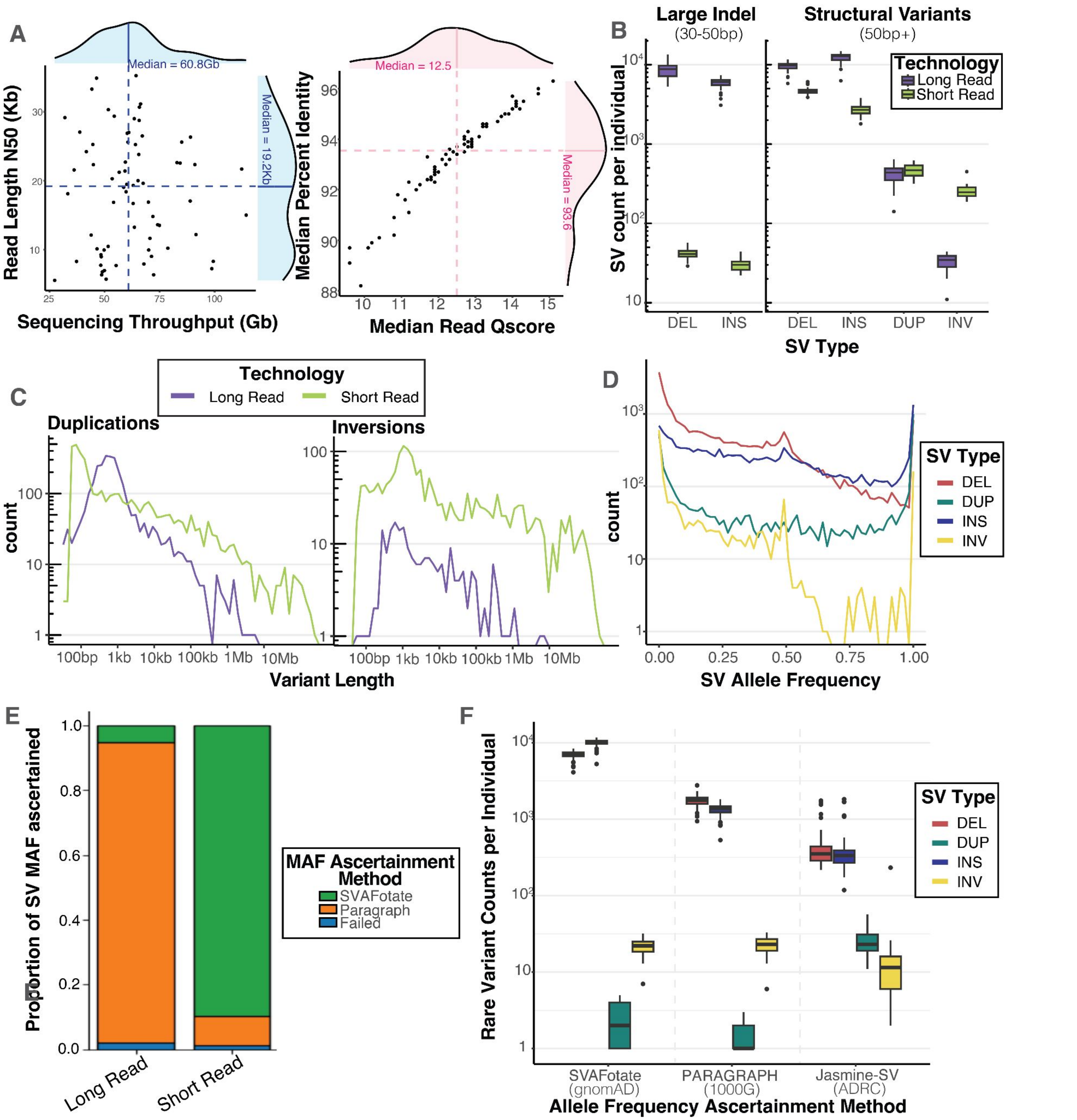


**Supplemental Figure 1 Sequencing statistics and SV characteristics**: **A.** Sequencing statistics from PomethION sequencing 68 individuals from the Undiagnosed Diseases Network. Total sequencing throughput in Gigabases is total yield of sequencing summed over all flowcells used for each sample. Read length N50 is a weighted median read length showing the read length at which most bases were sequenced in. After filtering low quality reads (Qscore < 7) and aligning to the GRCh38 reference genome, we display here the distribution of Read Quality scores and aligned median percent identity (what percent of the aligned reads matches perfectly to the reference sequence). **B** Counts of large indels and Structural variants detected by long-read and short-read technologies. Long-read variants were called with an ensemble algorithm merging individual callsets from sniffles, cuteSV, and SVIM. Short-read variants were called with Manta and then genotyped by PARAGRAPH. Structural variants are defined to be above 50bp whereas large indels span 30 to 50 bp. **C** Length and count distributions for Duplications and Inversions in 50 UDN samples who had both SRS WGS and LRS WGS. Large duplications and inversions up to 10 megabases were detected with short-reads, though these ultra-long events might be false positives. **D.** Allele frequency distribution of UDN structural variants called from SRS. Allele frequencies were ascertained by running SVAFotate and looking up variants in gnomAD, CCDG, and 1000G with a minimum overlap percentage of _% for matching. **E.** Comparison of different allele frequency ascertainment methods for both LRS-discovered and SRS-discovered variants. SVAFotate involves looking up in short-read reference populations (gnomAD, CCDG, 1000G) with an overlap percentage of _%. Paragraph involved running a local graph-based realignment algorithm to genotype breakpoints of SVs in SRS 1000G project genomes. Long–read shows very small ascertainment of variants in short-read population databases, and also had more variants that failed genotyping with paragraph. **F.** Counts of rare SVs detected from LRS per individual using different allele frequency ascertainment methods. As seen in F, SVAFotate poorly ascertains LRS-discovered variants, so filtering based on gnomAD MAFs results in a large number of variants present. Merging with ADRC, a technology-matched reference population, using jasmine-SV resulted in much more stringent filtering, easing burden of rare disease variant curation.

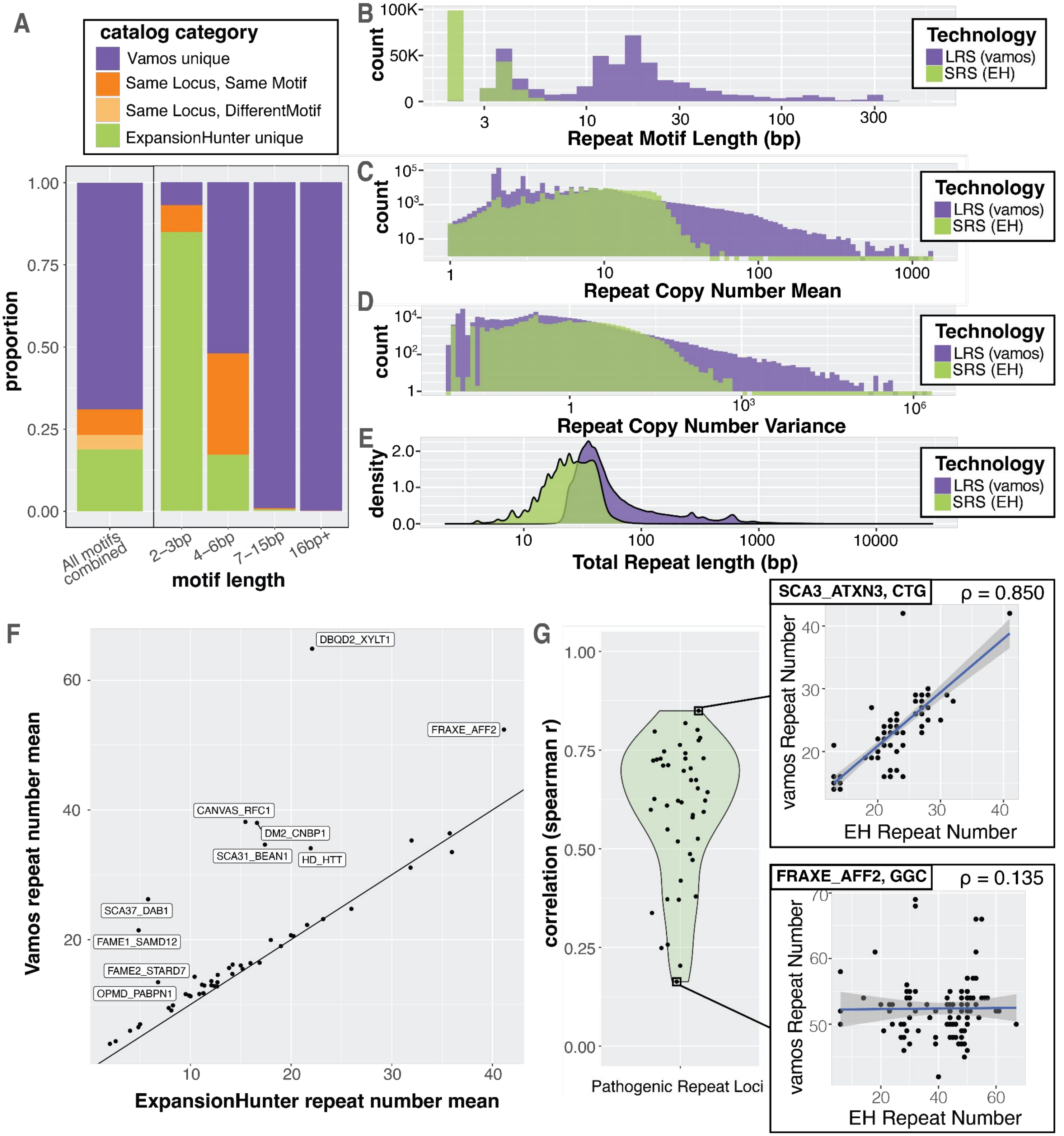


**Supplemental Figure 2 Long and short reads profile disease-relevant tandem repeat variation**. **A** Overlap of input catalogs to vamos (Long Reads) and ExpansionHunter (short reads), stratified by length of repeat motif. Vamos captures mainly long VNTRs > 7bp, while ExpansionHunters catalog is composed mainly of STRs (2-6bp). **B** Histogram of repeat motif length annotated in the respective catalog from vamos and ExpansionHunter (EH). **C** Histogram of the mean tandem repeat copy number across UDN genomes for each repeat in the vamos and EH catalog. Vamos profiles repeats that have larger mean length compared to EH. **D** Histogram of the variance of each tandem repeat copy number across UDN genomes. **E** Density of mean total repeat length in basepairs of the repeat across UDN genomes. Mean total repeat length is equivalent to mean tandem repeat copy number times the length of the repeat motif. **F** Dot plot showing correlation of tandem repeat copy number mean of 51 disease-causing repeat loci from STRchive as estimated by vamos and ExpansionHunter. Solid line showing the identity line y = x. Repeats with 10 biggest differences highlighted. **G** Distribution across all pathogenic repeats of the per allele correlation between vamos and ExpansionHunter estimated tandem repeat copy number. Correlation for the most correlated and least correlated repeat highlighted.

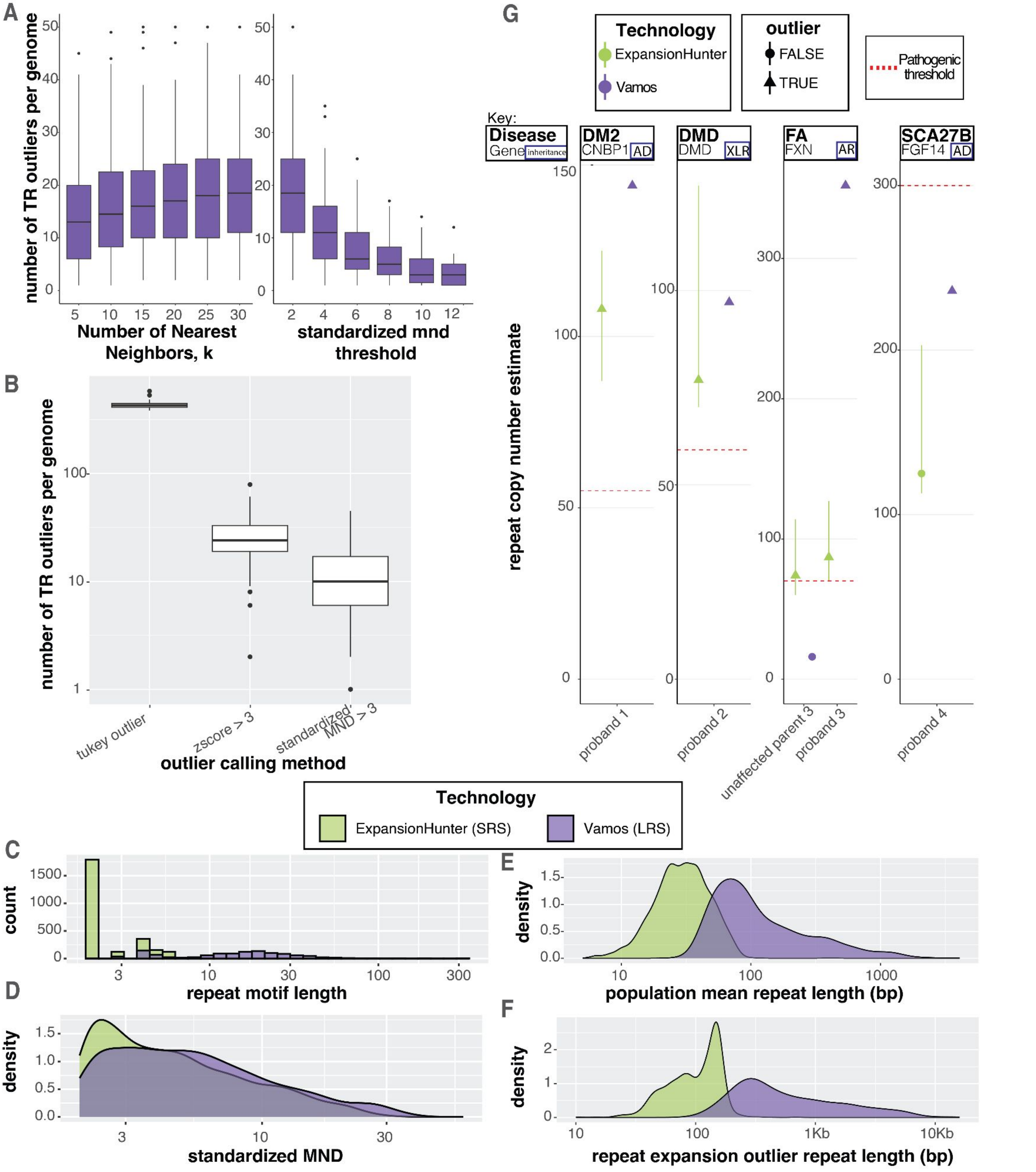


**Supplemental Figure 3 Extreme tandem repeat expansions detected from short and long-read sequencing. (A)** Varying parameters of mean neighbor distance (mnd) outlier method and its effect on number of extreme TREs detected per genome. Increasing k, number of nearest neighbors to calculate mnd, leads to more TREs per genome. While increasing the standardized mean neighbor distance (mnd) threshold decreases the number of TREs per genome. **B** Comparison of number of extreme TREs detected by different TR outlier methods. Tukey outlier calling outlier if sample is greater than 75% percentile + 1.5*IQR, and a standard z-score scaling method are compared with z threshold of 3 used to call outliers. MND methods results in less TRE calls per genome, triaging the most extreme examples. **C-F** Comparison of extreme TREs identified from Vamos with ONT long reads and with ExpansionHunter from Illumina short-reads. **C** histogram of the repeat unit length of vamos TRE outliers compared to ExpansionHunter outliers. **D** Standardized mean neighbor distance of vamos TRE outliers compared to ExpansionHunter outliers. **E** population mean repeat length across UDN individuals for the TRE outliers in vamos compared to ExpansionHunter. **F** Total length of the TRE outlier repeat in vamos compared to ExpansionHunter. **G** TRE outliers identified in STRchive pathogenic repeat loci in UDN individuals Pathogenic limit of the repeat labeled in red dotted line. Plotting only the length of the expanded allele.

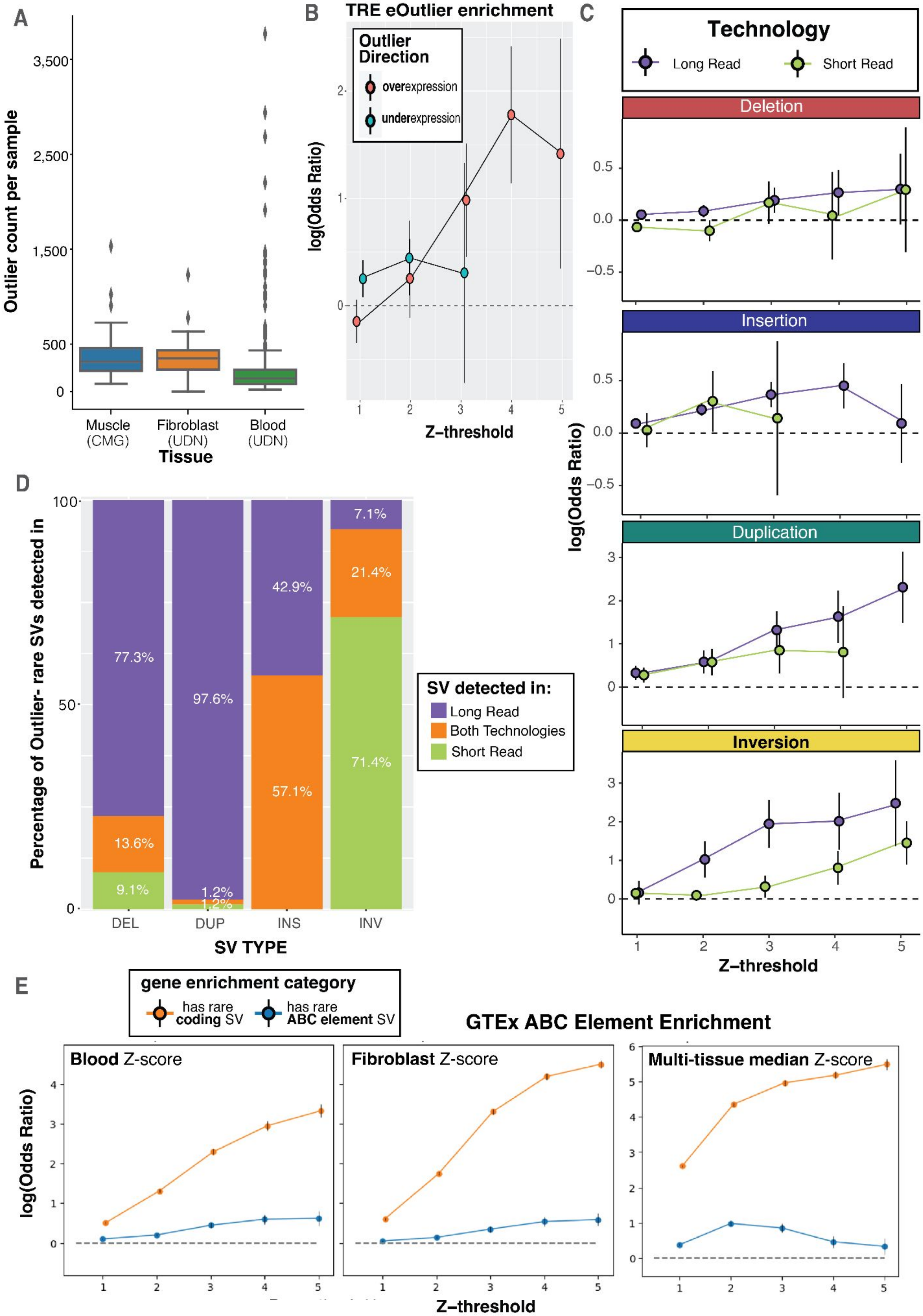


**Supplemental Figure 4. Rare SV enrichments from both short and long reads nearby expression outliers. A** Count of expression outliers (absolute z-score > 3) per individual across tissues after normalizing and residualizing RNAseq data. **B** Enrichment of extreme TRE outliers nearby either over or under expression outliers at varying z-thresholds. Model did not converge for underexpression outliers above z=3 due to lack of examples. **C** Comparison of enrichments from long-read vs short-read discovered SVs from a subset of individuals with short-read WGS data available. Data absent where model did not converge due to a lack of rare SV-outlier examples. **D** Percentage of rare SVs found nearby expression outliers that were detected from either technology. **E** Enrichment of SVs overlapping ABC elements for expression outliers of their target genes, controlling for enrichment of coding SVs across different expression z-scores. Single tissue expression z-scores displayed modest enrichment of disrupting ABC enhancers, while the multi-tissue median z-score did not.

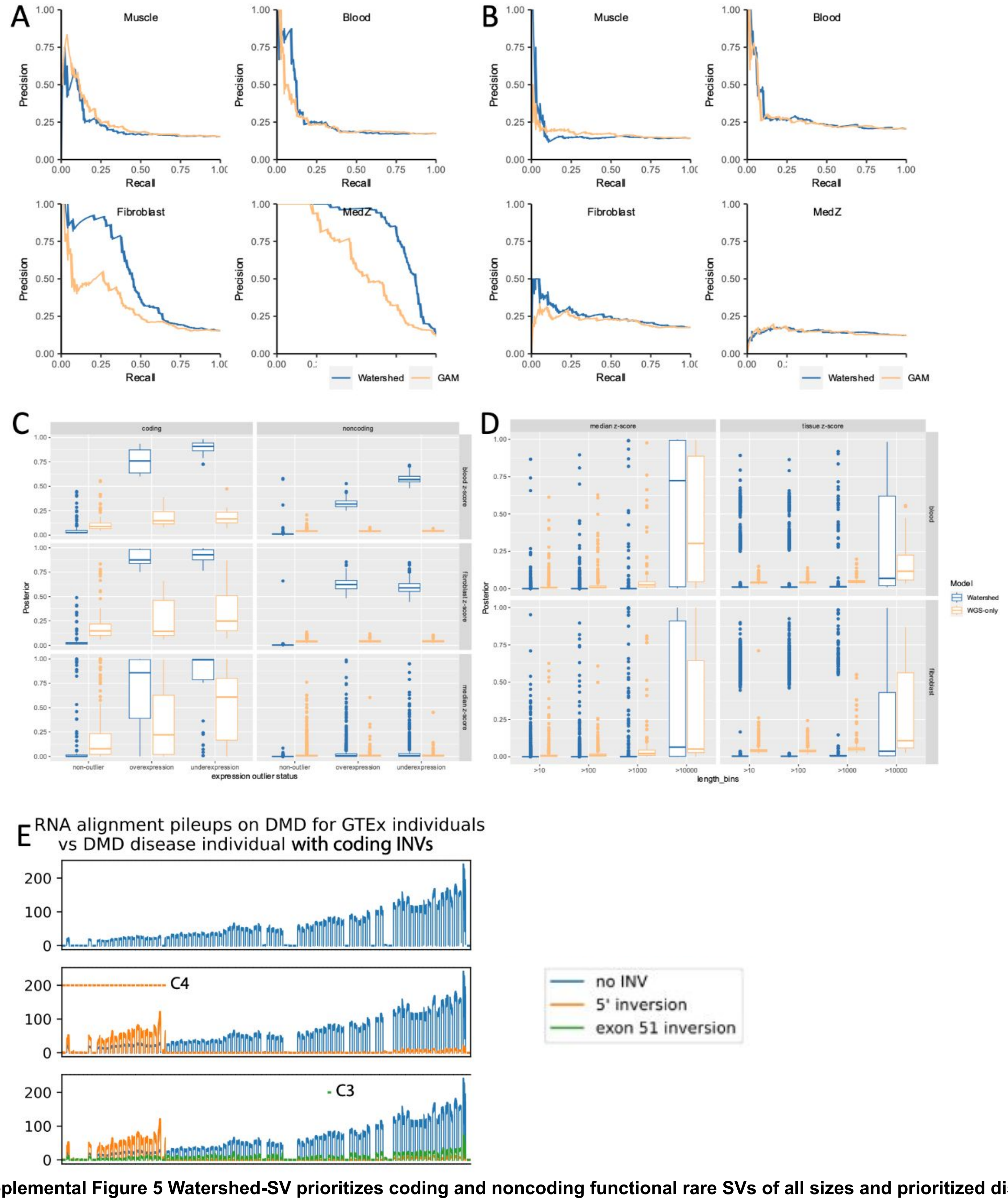


**Supplemental Figure 5 Watershed-SV prioritizes coding and noncoding functional rare SVs of all sizes and prioritized disease SV. (A)** Precision-Recall Curves (PRC) of benchmark using held-out N2 pairs in 3 single-tissue and 1 multi-tissue Watershed-SV models at 100kb distance limit against WGS-only model with the same setup. **(B)** Precision-Recall Curves (PRC) of benchmark using held-out noncoding N2 pairs in 3 single-tissue and 1 multi-tissue Watershed-SV models at 100kb distance limit against WGS-only model with the same setup.**(C)** Using a z-score threshold of -3 and 3, we stratified 100kb multi-tissue, blood, fibroblast per-tissue Watershed-SV models' predictions on UDN dataset posterior probabilities by under-, over-, and non-outliers (categories), and plotted by coding vs noncoding variants (column), and which model is used to predict the respective RNA tissue type; each dot represent an gene-SV pair. **(D)** Posterior probability distribution of Watershed-SV vs WGS-only model stratified by length of SV. Grid column represents model used (multi-tissue Watershed-SV (Left), per-tissue Watershed-SV (Right)). **(E)** All 2 DMD patients with diagnostic rare SVs from CMG data are prioritized by Watershed-SV, top row is GTEx control's mean RNA-depth across exons of DMD gene. Two bottom rows represent adding one patient at a time. Bars hovering above piles are indicator of being affected by diagnostic rare SV.

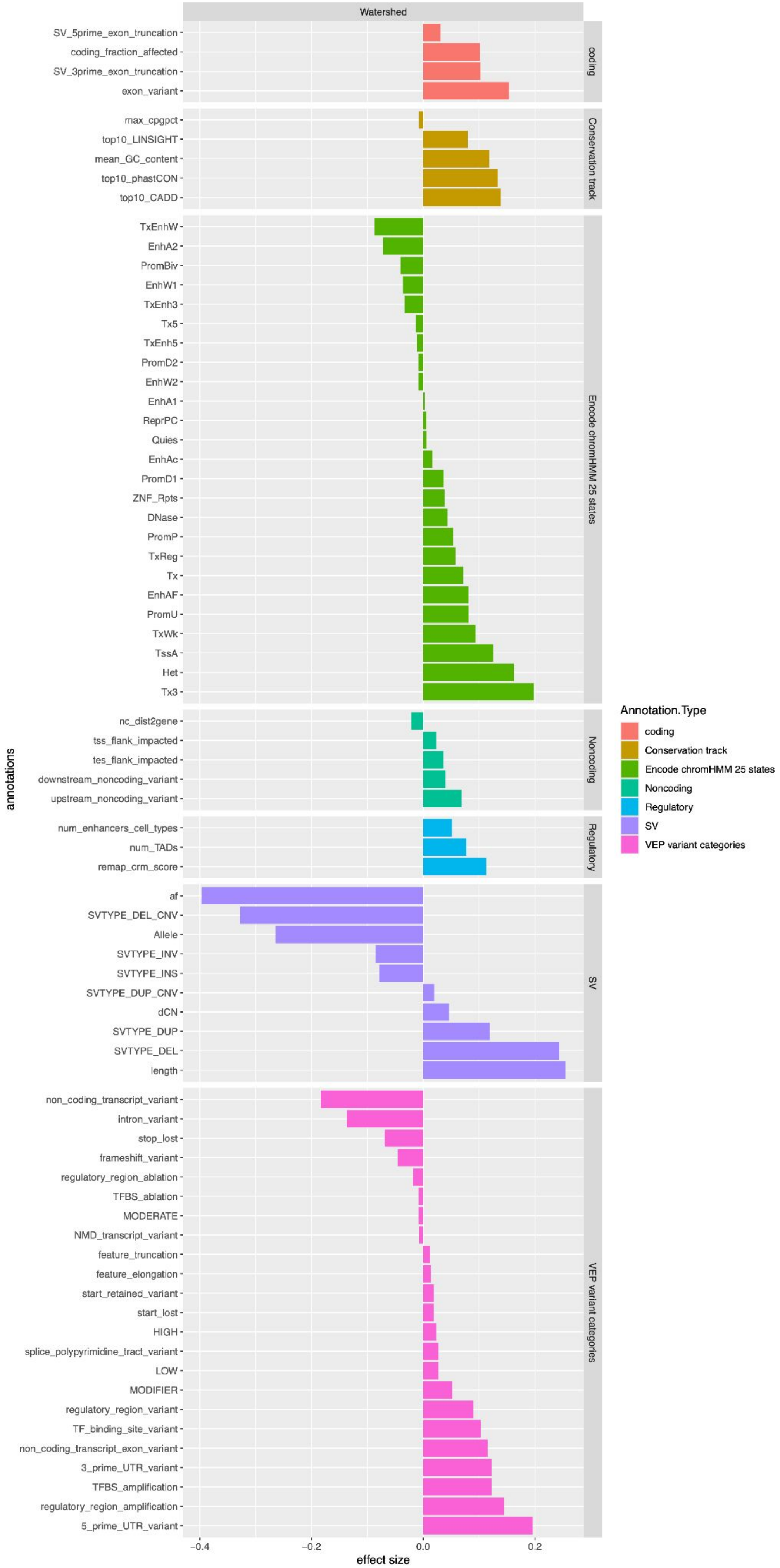


**Supplemental Figure 6 10kb multi-tissue Watershed-SV model effect-sizes.** Effect sizes are segregated into categories.

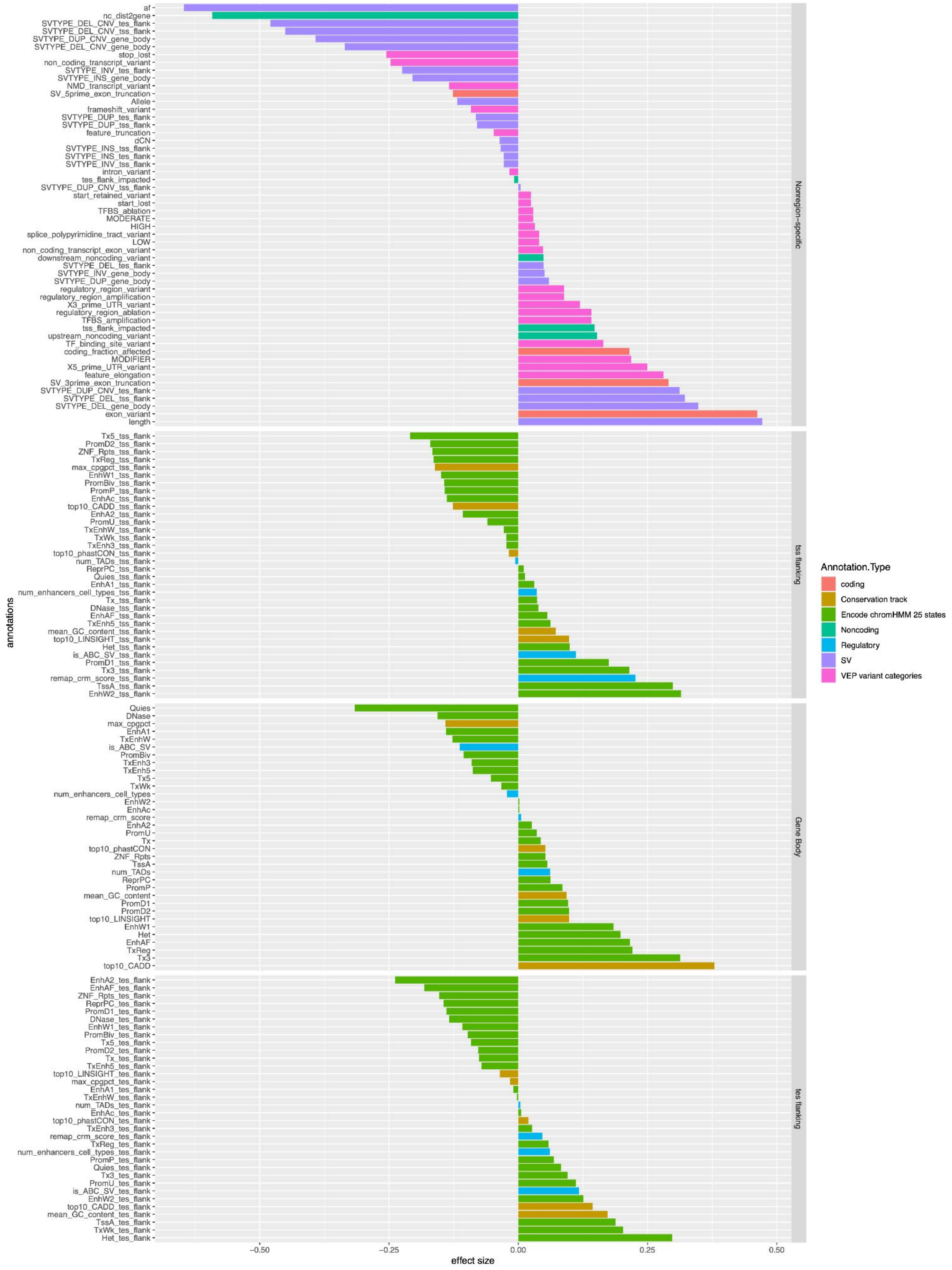


**Supplemental Figure 7 100kb multi-tissue Watershed-SV model effect-sizes.** Effect sizes are segregated into categories and then by region if they are ChromHMM annotations, conservation scores, or regulatory annotations.

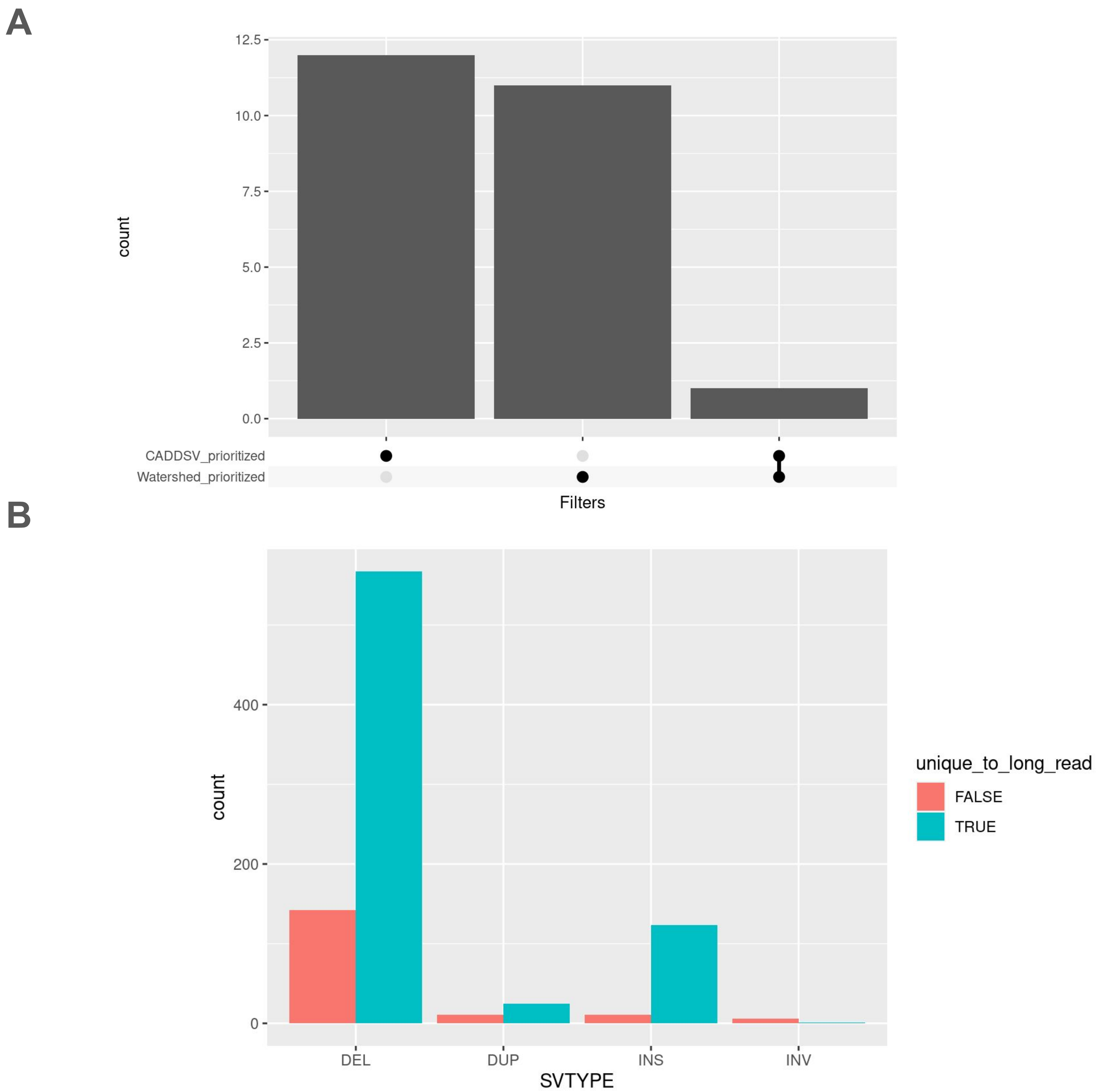


**Supplemental Figure 8 Watershed-SV prioritizes additional rare LRS SVs relevant to patient HPO terms (A)** Upset plot of number of gene-SV pairs from UDN LRS dataset prioritized by either Watershed-SV(>60%) or CADD-SV (>10) with Phen2Gene Rank < 300. (B) Watershed-SV prioritized UDN LRS gene-SV pairs by whether they're unique to LRS discovery or discovered by both LRS and SRS. Grouped by SV types.

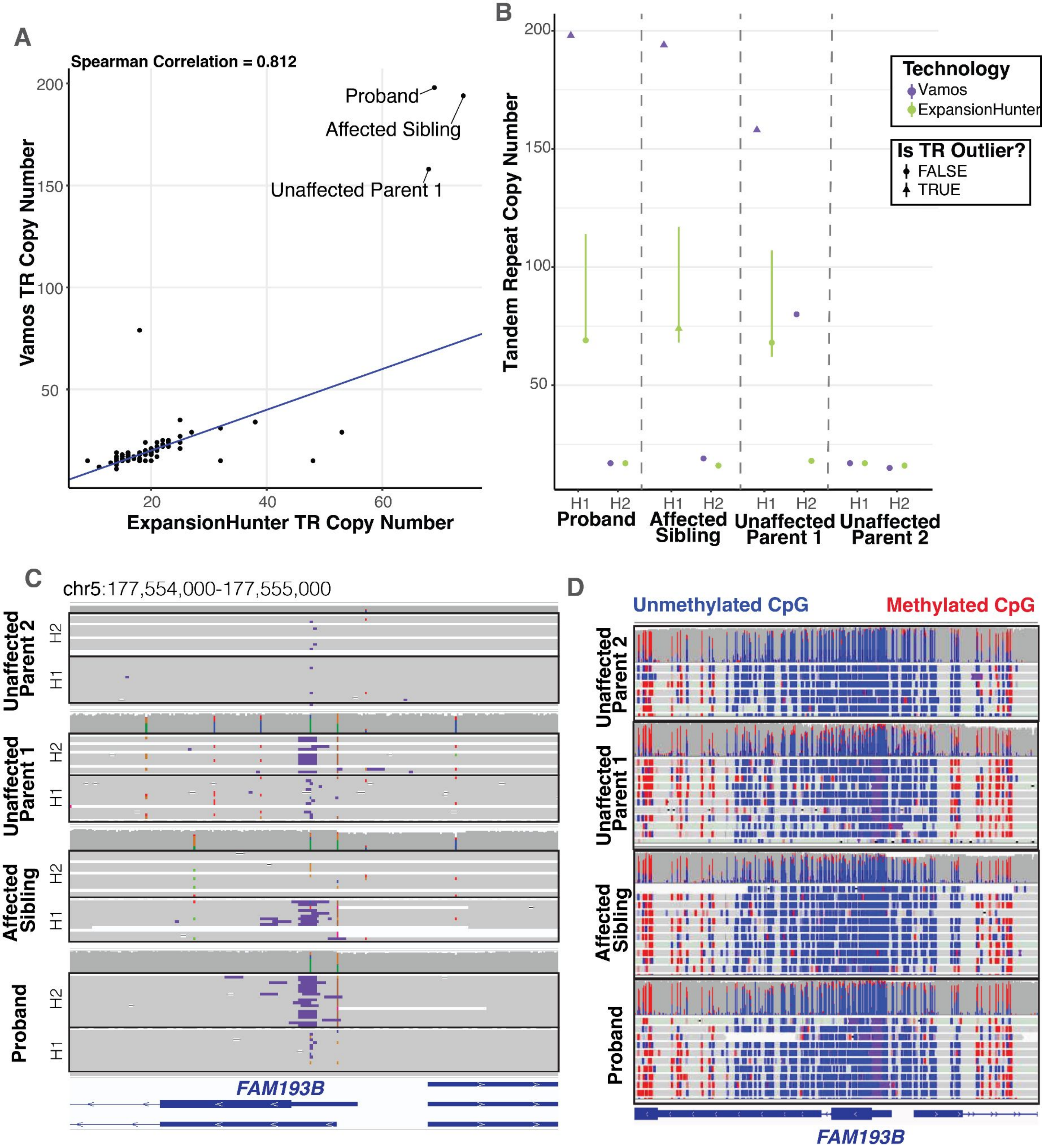


**Supplemental Figure 9 Extreme Tandem Repeat Expansion at Candidate Disease Locus. A** Correlation of allele specific tandem repeat copy number at the FAM193B STR estimated by vamos and ExpansionHunter across UDN cohort. Blue line showing identity line y = x. **B** For family with the extreme tandem repeat expansion, comparison of tandem repeat copy number in each haplotype by technology. For consistent comparison, H1 is assigned to allele with largest copy number and H2 assigned to allele with smallest. **C** Screenshot of phased IGV pileup of the FAM193B 5' UTR showing insertions as purple bar within each read. **D** Screenshot of IGV pileup of the zoomed out promoter / CpG island around the TRE colored by methylation. Blue represents CpGs that were predicted to be unmethylated, red represents those predicted to be methylated. No change in methylation observed in the affected carriers of the expansion compared to the unaffected parents.
